# Ethnic inequalities in respiratory virus epidemics in England: a mathematical modelling study

**DOI:** 10.64898/2026.04.18.26350858

**Authors:** Alexis Robert, Lucy Goodfellow, Lorenzo Pellis, Edwin van Leeuwen, W John Edmunds, Billy Quilty, Kevin van Zandvoort, Rosalind M Eggo

## Abstract

**Background:** In England, the burden of respiratory infections varies by ethnicity, contributing to health inequalities, but the role of additional demographic factors remains underexplored. We quantified how differences in social mixing and demographic characteristics between ethnic groups cause inequalities in transmission dynamics.

**Methods:** We analysed the association between the ethnicity and the number of contacts of 12,484 participants in the 2024–2025 Reconnect social contact survey, using a negative binomial regression model. We simulated respiratory pathogen epidemics using a compartmental model stratified by age, ethnicity, and contact levels, at a national level and in major cities in England.

**Findings:** After adjusting for demographic variables, participants of Black and Mixed ethnicities had more contacts than those of White ethnicity (rate ratios (RR): 1.18 [95% Credible Interval (CI): 1.11-1.26], and 1.31 [95% CI: 1.14-1.52]). Participants of Asian ethnicity had fewer contacts (RR: 0.85 [95% CI: 0.79-0.91]). In national-level simulations, individuals of White ethnicity had the lowest attack rates due to demographic differences and mixing patterns. Local demographic structures changed simulated dynamics: attack rates in individuals of Black and Mixed ethnicities were approximately double those of White ethnicity in Birmingham, but less than 60% higher in Liverpool.

**Interpretation:** Demographic characteristics and mixing patterns create inequalities in transmission dynamics between ethnicities, while local demographic characteristics and pathogen infectiousness change the expected relative burden. To ensure mitigation strategies are effective and equitable, their evaluation must explicitly account for inequalities arising from local context.

**Funding:** Medical Research Council, National Institute for Health and Care Research, Wellcome Trust

**Research in context:** 

**Evidence before this study:** We searched PubMed for population-based studies quantifying differences in respiratory infections between ethnic groups, up to 1 April 2026, with no language restrictions. Keywords included: (respiratory pathogens OR influenza OR COVID-19) AND (ethnic* OR race) AND (inequ*) AND (compartmental model OR incidence rate ratio OR hazard ratio). We excluded studies that focused on non-respiratory pathogens (e.g. looking at consequences of COVID-19 on incidence of other pathogens). A population-based cohort study showed that influenza infection risk was higher in South Asian, Black, and Mixed ethnic groups compared to White ethnicity in England. Another population-based cohort study highlighted that during the first wave of COVID-19 in England, the South Asian, Black, and Mixed ethnic groups were more likely to test positive and to be hospitalised than the White ethnic group. Census data in England showed that the distributions of age, household size, household income and employment status differed between ethnic groups, and the recent Reconnect social contact surveys highlighted the impact of each demographic factor on the participants’ number of contacts.

**Added value of this study:** Our study shows that social contact patterns, mixing, and demographic structure all lead to unequal infection risk between ethnic groups in respiratory pathogen epidemics. Using the largest available social contact survey in England, we show that both the average number of contacts and the proportion of high-contact individuals varied by ethnic group, even after adjusting for participants’ demographics. These differences, together with mixing patterns and age structure, led to lower expected incidence among individuals of White ethnicity than in all other ethnic groups in simulated outbreaks. The level of inequality between ethnic groups changed when we used different values of pathogen transmissibility. Finally, as ethnic composition and population structure differ between cities in England, our results show differences in expected inequalities at a local level.

**Implications of all the available evidence:** Inequalities in infection risk between ethnic groups are context- and pathogen-dependent. They arise from both local population structure and contact patterns. Detailed information on mixing between groups and population structure is needed to accurately measure group-specific infection risk. These findings indicate that public health interventions based only on national-level estimates conceal regional variation in risk and may ultimately increase inequalities. Public health interventions need to be tailored to local contexts to be equitable and effective. Finally, our findings provide a foundation for understanding the progression from infection-risk inequalities to disparities in disease presentation and clinical outcomes.

## Introduction

The burden of infectious diseases is unequal: in the United Kingdom, individuals from minoritised ethnic groups and living in deprived areas are at higher risk of infection (1,2), hospitalisation, and severe outcomes (3,4) across respiratory pathogens. Inequalities arise from multiple factors, both demographic and structural, including access to healthcare (5), age distribution, social behaviour associated with occupation or cultural norms (6), household size and composition, and living conditions (7,8). To develop effective and equitable public health interventions, we need to better understand what drives differences in transmission dynamics of respiratory pathogens.

The distribution of social contacts is known to affect transmission dynamics for respiratory pathogens. Individuals with a high number of contacts have higher risks of being infected, and of transmitting the infection to others (9). Furthermore, individuals of different social groups do not mix homogeneously: intragroup mixing is typically much more frequent than intergroup mixing (10), affecting transmission between social groups. Finally, the distribution of contacts is dispersed, with some individuals having a much higher or lower number of contacts than the average. The average number of contacts, the assortativity of contacts, and the dispersion of the contact distribution are all highly correlated with demographic and socioeconomic variables, which differ by ethnicity: in England, individuals identifying as White British are on average older and live in less deprived areas (11,12). Differences in demographic characteristics between ethnic groups will therefore create expected differences in attack rates.

The Reconnect survey was a nationally representative social contact study carried out in 2024-2025 in the United Kingdom, which collected the characteristics of participants and their daily contacts including age and ethnicity (13). We analysed the association between the participants’ ethnic groups and their distribution of contacts, adjusting for demographic characteristics. We then estimated the number of infections expected in each ethnic group given the distribution of contacts and social mixing patterns using a mathematical model stratified by age, ethnic group, and level of contact. Finally, we quantified how differences in demographic characteristics between cities impact local epidemic dynamics generated by the model.

## Methods and Data

A summary of the analysis plan showing datasets and methods used in the analysis is available in the Supplementary Material (Figure S1).

### Social contact data

Reconnect was a cross-sectional survey in which social contact data were collected from 13,238 participants in the United Kingdom between November 2024 and March 2025 (13). For each participant, data including their age, gender, ethnicity, household size, residence, occupation type and household income were collected, in addition to their total number of contacts (including all individual and large group contacts) in the 24 hours preceding the survey and importantly, characteristics of their contacts, including age, sex, ethnicity, and day of contact. Ethnicity was self-reported, using the 2021 census categories (Asian, Black, Mixed, Other, and White). Participant residence was classified as urban or rural based on the first half of their postcode. After excluding participants with missing entries on necessary variables, the final dataset contained 12,484 participants.The distribution of variables is shown in Supplementary Section S2.

### Demographic data

We extracted the distribution of demographic variables in different areas: England (country level), London (regional level), Birmingham, Leicester, Liverpool, Manchester, and York (lower tier local authority level, a smaller scale than the regional level). Distributions of household size and economic activity status by age and ethnic group were taken from the Census 2021 data for England and available for all geographical units (12). Stratification of economic activity status by age, ethnic group and sex was available at a national level, but only the stratification by age and ethnic group was available at local levels due to the Census’ data protection requirements. We approximated the distribution of employment status by sex at a local level using the national-level distribution. We classified all economically active and in employment individuals (part-time or full-time) as Employed, and all individuals classified as economically active and unemployed as Unemployed. We re-labeled “economic activity status” as “employment status” to match the label used in the Reconnect survey. The weekly household income by ethnicity was taken from the Family Resources Survey in the United Kingdom (14). Only the national-level distribution was available.

In Reconnect, participants were asked their gender, and were asked to report the sex of their contacts. The 2021 census data recorded sex in two categories (“Male”, “Female”). We use ‘gender’ for simplicity throughout the analysis.

### Bayesian negative binomial regression analysis

We used Bayesian negative binomial regression to analyse how the mean and dispersion of the number of contacts per participant in the Reconnect study were associated with ethnicity. We used a negative binomial model to account for differences in mean and in dispersion, where larger values of the dispersion parameter indicate less dispersed distributions. The regression model was implemented using the R package brms (15). The model was run with four Markov chains, each with 2,000 iterations and no thinning, including a warmup period of 1,000 iterations.

In Reconnect, 92.8% of participants from rural areas identified their ethnic group as White, in line with the distribution of ethnicity in urban and rural areas in England (12). We created a composite variable merging ethnic groups and urban/rural status (Supplementary Section S2). We adjusted for the impact of variables collected in Reconnect on the average number of contacts: participant’s age group, employment status, annual household income, household size, gender, and whether the survey day was a weekday. All model parameters were set up with a uniform prior distribution (Supplementary Section S3).

### Compartmental model stratified by age, ethnicity, and contact group

To explore the impact of differences in mean and dispersion in number of contacts on epidemic dynamics, we implemented a stochastic compartmental transmission model with susceptible, exposed, infected and recovered compartments (SEIR model). The model was stratified by age group, ethnicity, and by levels of contact group to capture the dispersion of contact distribution in each age and ethnic group.

We used the parameter estimates from the regression analysis to classify individuals in the population into three contact groups. For each stratum of age and ethnic group, we computed the mean number of contacts of low, medium, and high contact individuals, and the proportion of individuals in each contact group. To do so, we simulated the contact distribution separately in each age–ethnic stratum: We generated 500 synthetic individuals per age-ethnicity stratum, and assigned each a household size, employment status, gender, and household income using the distribution by age and ethnic group in the population (Supplementary Section S4). We simulated the number of contacts for each individual using five parameter draws from the regression outputs, leading to 2,500 simulated contact counts per age-ethnicity stratum. We simulated the number of contacts using the “urban” and “weekday” levels for each individual. We then fitted a three-component Poisson mixture model to the simulated contact counts in each age-ethnicity stratum to classify individuals into low-, medium-, and high-contact groups. The parameters of the mixture model correspond to the mean number of contacts in each group, and the proportion of individuals belonging to that group.

In order to capture the assortativity in mixing between social groups the compartmental model used a mixing matrix between all age, ethnicity and contact group strata. To compute the mixing matrix, we combined the separate age and ethnicity transmission matrices from the Reconnect survey (13), assuming that ethnicity mixing was the same across age groups (and vice versa) and did not depend on the age distribution per ethnicity. We used the number of contacts per contact group to compute the age-ethnicity-contact group mixing matrix. See Supplementary Section S5 for details on the calculation of the overall contact matrix.

### Stochastic simulations and scenarios

We simulated an influenza-like epidemic (incubation period of 3 days, infectious period of 5 days), starting with 30 infected individuals with no vaccination or waning of immunity. The transmission model was coded in R, using the packages odin2 and dust2 (16) and code is available at https://github.com/alxsrobert/connect_overdispersion. We set the transmission rate, *β*, to get *R_0_* ranging from 1.4 to 8.0, a range that contains the basic reproduction number of most common respiratory pathogens. The model was initially run using the population size and the age and ethnicity distribution from the 2021 Census for England (12).

In order to separately understand the effect of i) demographic differences, ii) social mixing patterns, and iii) regression parameters on infection incidence by ethnic group we implemented three alternative scenarios. First, to observe the differences due to demographic structure and mixing patterns alone, we set all ethnicity-related regression parameters to 1 (i.e. adjusting for demographic variables, all ethnic groups had the same distribution of contacts). This changed the distribution of contact groups per age and ethnicity stratum. Second, to quantify the heterogeneity due to regression parameters and mixing patterns, we used the national-level distribution of demographic variables (age, household size, income and employment status) in all ethnic groups. This changed the distribution of age and contact groups per ethnicity. Thirdly, we ran simulations using the population-level distribution of demographic variables, and homogeneous mixing between ethnic groups (i.e. we replaced the per capita mixing matrix between ethnicities from Reconnect with a constant matrix). This changed the distribution of age, contact groups per ethnicity, and mixing matrix in the model, and the only remaining source of heterogeneity between ethnic groups was the ethnicity-related parameters from the regression.

To determine how differences in ethnicity and demography in major cities in England could affect epidemics and inequalities, we generated stochastic simulations using the local distribution of demographic characteristics in six urban centres in England: Birmingham, Leicester, Liverpool, London, Manchester, and York. Using local distributions changed the overall population size, and the distribution of age, ethnicity, and contact groups. We did not re-fit the regression analysis or recompute the per capita matrices. We also generated simulations keeping *β* fixed across different cities to evaluate how changes in the population structure resulted in local differences in *R_0_* and attack rate.

There were two sources of stochasticity in the simulation process: the classification into contact groups (resulting from the synthetic population and sampling from the regression outputs), and the stochastic SEIR model. To account for both, we generated 100 different synthetic populations, and ran 15 simulations per population, leading to 1,500 stochastic simulations per value of *R_0_* and per scenario.

## Results

### Comparison of demography and contact distribution between ethnicities in England

In England in 2021, White individuals tended to be older (median age: 43 years old), while individuals of Mixed ethnicities were younger (median age: 18 years old) (12) (Figure 1, Panel A). The age distribution affected the distribution of employment status: 24.8% of individuals of White ethnicity were retired, compared to 4.8% to 9.0% in all other ethnic groups (Figure 1, Panel D) (12). Individuals from the White ethnic group tended to live in smaller households (45.8% in households of two people or fewer) (Figure 1, Panel C) (12). The smallest proportion of households with an annual income above £100,000 was observed in Black households (4.0%).

**Figure 1:**
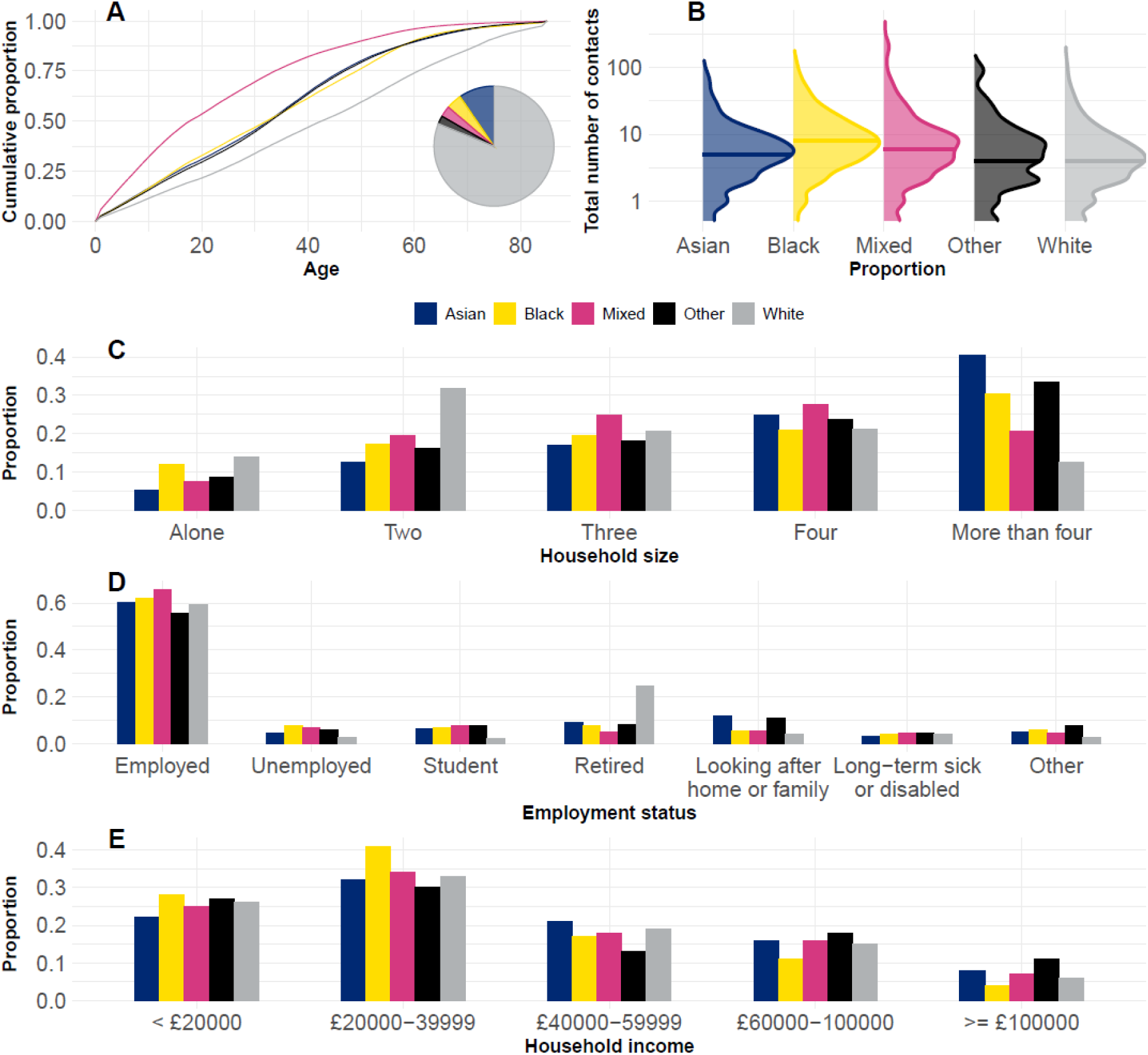
Demographic characteristics of the population of England. A. Cumulative age distribution of the population of England, stratified by ethnicity, with the pie chart insert showing the distribution by ethnicity in England (from 2021 Census data (12)). B. Distribution of the number of contacts by ethnicity (from Reconnect (13)). Distribution of household size (C) and employment status (D) by ethnicity in England (from 2021 Census data (12)). The “student” level refers to individuals classified as students and not economically active. E. Distribution of annual household income by ethnicity in the United Kingdom (computed from the Family Resources Survey (14)). In Reconnect, categories of annual incomes were reported. We matched the weekly and annual income categories by multiplying the boundaries of the categories of weekly income by 50 (if we had multiplied by 52, values would have straddled the annual income categories defined in Reconnect). According to the Family Resources Survey, median annual household income in the United Kingdom was between £30,000 and £40,000.

In the Reconnect survey, among the 12,484 participants considered here, 79.6% identified their ethnic group as White, 9.4% Black and 7.9% Asian, 2.5% Mixed, and 0.6% Other. The median number of contacts was highest in individuals of Black ethnicity (8 contacts), and lowest in individuals of White ethnicity (4 contacts) (16). The proportion of individuals who reported more than 100 contacts was highest in participants of Mixed ethnicity (2.6%, below 1% in all other ethnic groups).

### Differences in number of contacts between ethnicities remain after controlling for demographic factors

Ethnicity was associated with differences in the mean and dispersion of the number of contacts after adjusting for demographic characteristics (Figure 2). On average, in urban settings, individuals of Asian ethnicity had fewer contacts than White individuals (RR: : 0.85 [95% CI: 0.79-0.91]). Individuals of Black ethnicity had more contacts than White individuals (RR: 1.18 [95% CI: 1.11-1.26]), and so did individuals of Mixed ethnicity (RR: 1.31 [95% CI: 1.14-1.52]). Individuals of White ethnicity living in rural areas were associated with a slightly lower average number of contacts than individuals from urban areas (RR: 0.95 [95% CI: 0.89-1.01]).

**Figure 2:**
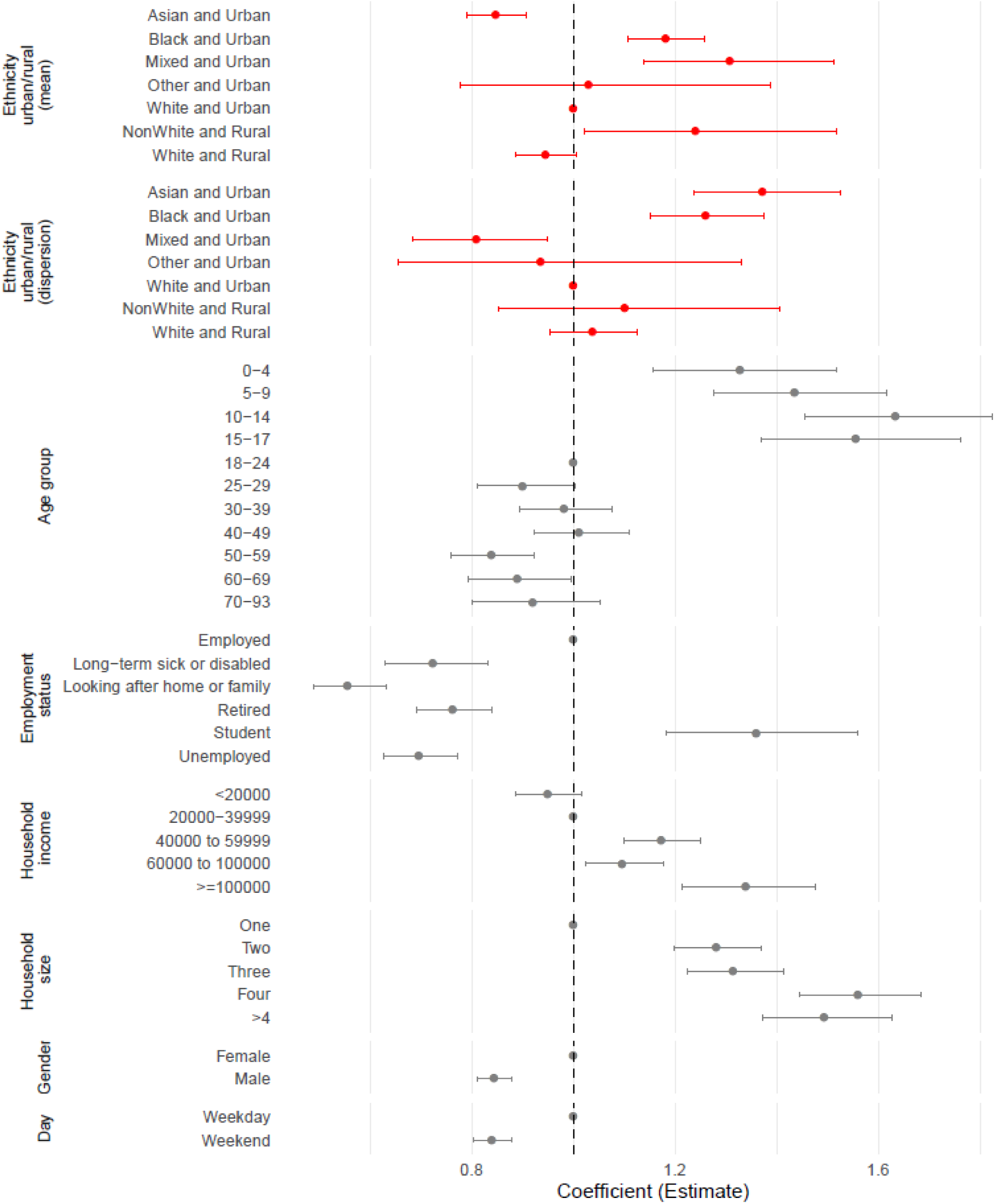
Parameter estimates (median and 95% credible intervals of the rate ratios) from the Bayesian Negative Binomial regression analysis on the number of social contacts. Parameter estimates are also shown in Supplementary Section S6. Due to wide credible intervals, parameters estimates for Other gender and Other employment status are not shown in this figure. Red rows highlight ethnicity-related parameters, all other rows are referred to as “demographic variables”. The “ethnicity urban/rural (dispersion)” parameters are the only parameters affecting the dispersion, all other parameters only affect the mean. High values of dispersion parameters indicate less dispersed distributions.

In urban settings, the distribution of contacts in individuals from Asian and Black ethnic groups was less dispersed than in White individuals (RR: 1.37 [95% CI: 1.24-1.51] and 1.26 [95% CI: 1.15-1.38] respectively), while the distribution of contacts was more dispersed in individuals of Mixed ethnicity (RR: 0.81 [95% CI: 0.69-0.94]), indicating a higher proportion of individuals with extreme contact numbers. There was no statistically significant difference in the dispersion of contacts between White individuals and individuals of Other ethnicities.

Children and teenagers had on average more contacts (RR: 1.44 [95% CI: 1.28-1.61] in 5-9 year olds, 1.56 [95% CI: 1.37-1.77] in 15-17 year olds) than 18-24 year olds. Student adults were associated with more contacts than employed adults (RR: 1.36 [95% CI: 1.19-1.56]), while retired individuals had much fewer contacts (RR: 0.76 [95% CI: 0.69-0.84]). Average number of contacts increased with household size but plateaued between households of size four and larger households (RR: 1.56 [95% CI: 1.44-1.69] when household size was 4, 1.49 [95% CI: 1.37-1.63] when household size exceeded 4). The average number of contacts increased with household income. Parameter estimates are shown in Supplementary Section S6.

The regression coefficients and ethnicity-specific demography led to differences in simulated contact distribution (Supplementary Section S7): Individuals of Asian and White ethnicity had a similar mean number of contacts (9.1 [95% Simulation Interval: 8.6-9.5] and 9.2 [95% SI: 8.8-9.6] respectively), while individuals of Black ethnicity had more contacts on average (12.1 [95% SI: 11.7-12.6]).

### Infection inequalities were generated by ethnicity-specific number of contacts, demographic characteristics, and mixing patterns

Across all values of *R_0_*, the attack rate in simulated outbreaks was lowest in the White ethnic group (Figure 3A, first column). The attack rate was highest in the Mixed (relative attack rate compared to the White ethnic group: 1.49 [95% SI: 1.44-1.54] at *R_0_* = 2) and Black (1.41 [95% SI: 1.38-1.44] at *R_0_* = 2) ethnic groups. The relative attack rate in the Asian ethnic group was roughly constant, between 1.05 and 1.08 across the *R_0_* range. For all values of *R_0_*, the age-standardised relative attack rate was highest in individuals of Black ethnicity (Supplementary Section S8). The majority of cases were from the White ethnic group (Supplementary Section S9), as the group constitutes almost 80% of the population in England. High contact individuals were at higher risk of becoming infected, and the attack rate decreased with age (Supplementary Section S9).

**Figure 3:**
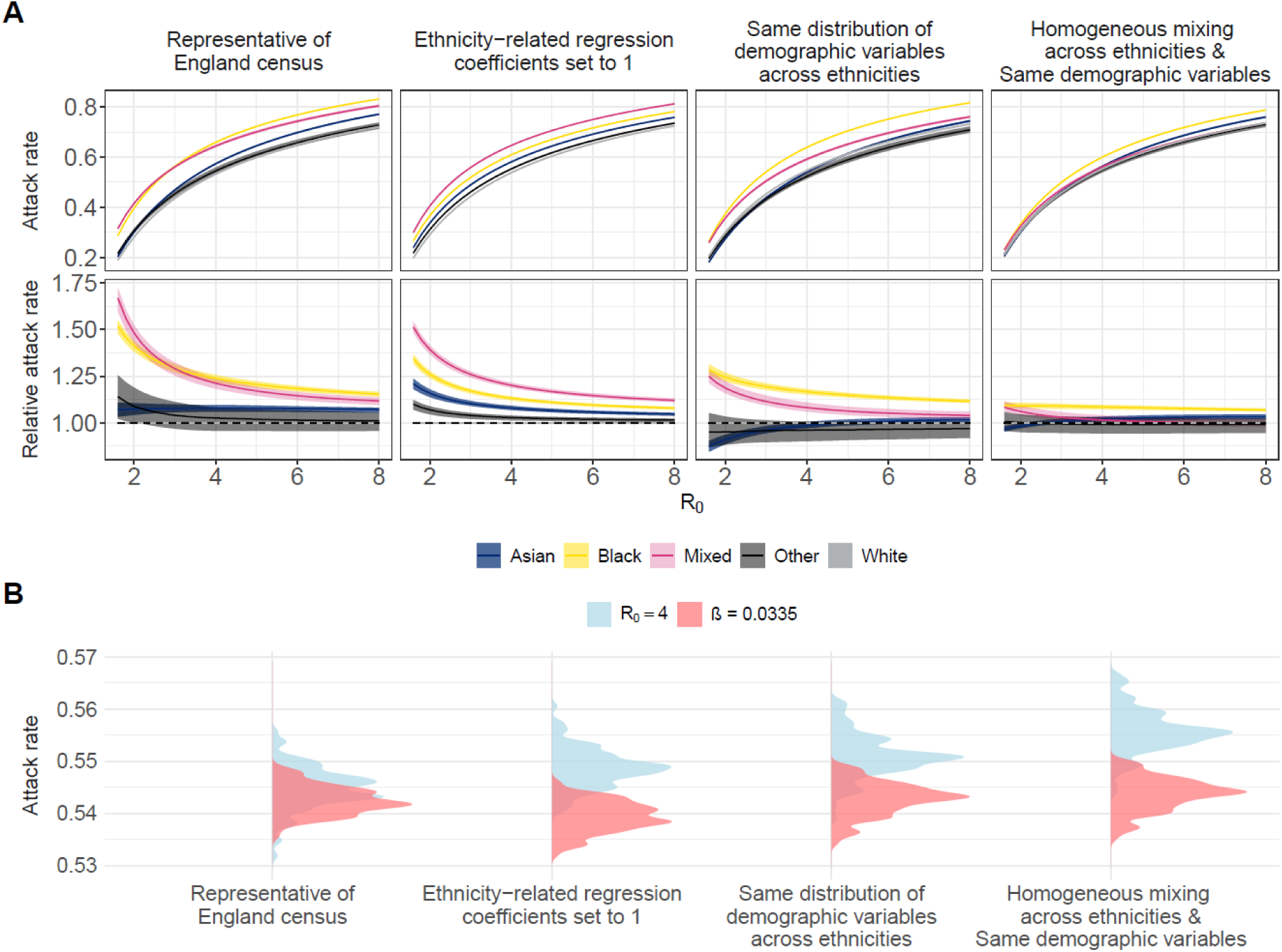
Simulated transmission dynamics and sources of heterogeneity. A. Top panels: Proportion of each ethnic group infected across the course of the epidemic for each scenario and value of R*_0_*. A. bottom panels: Relative attack rate compared to the White ethnic group for each scenario and value of R*_0_*. The lines represent the median values, and the ribbons show the 95% Simulation intervals. B. Density of the overall proportion of the population infected for each scenario, adjusting *β* to keep R*_0_* = 4 constant in all four scenarios or keeping constant *β* = 0.0335 (which corresponds to R*_0_* ∼4 in the “Representative of England census” scenario). Keeping *β* constant instead of R*_0_* led to a similar attack rate for all scenarios, while keeping R*_0_* constant led to slight changes in attack rate.

When differences between ethnic groups were due to differences in demographic structure and mixing patterns between ethnic groups alone, heterogeneity in attack rates decreased. In particular in Black and Mixed ethnicities (Figure 3A, second column), for whom the mean number of contacts in the regression analysis was highest. Setting the distribution of demographic variables per ethnic group to the national-level distribution greatly decreased the heterogeneity between attack rates (Figure 3A, third column), especially in the Mixed ethnic group (Maximum median relative attack rate of 1.25, compared to 1.68 in the reference set of simulations). The remaining heterogeneity was caused by differences in contact distribution and mixing patterns between ethnic groups. Finally, when heterogeneity only stemmed from differences in contact distributions from the regression outputs, some differences in attack rates remained, but were much lower than in all previous scenarios (Figure 3A, fourth column): Across values of *R_0_*, the relative attack rate was highest in the Black ethnic group (1.10 [95% SI: 1.08-1.11] at *R_0_* = 2).

When *R_0_* was held constant, the overall attack rate increased when we removed heterogeneity in contact rate between ethnicities, but if the transmissibility rate, *β*, was held constant the attack rate was approximately the same in all scenarios (Figure 3B). We tested if the number of contact groups affected results, and using four contact groups instead of three did not change the simulation outputs (Supplementary Section S10). The added heterogeneity led to a slight decrease in overall attack rate (from 54.5% [95% SI: 54.1-55.0] to 53.0% [95% SI: 52.1.1-53.8] at *R_0_* = 4), but the relative attack rates by ethnic group were similar to the main scenario.

### City-specific distribution of demographic variables impacts incidence and *R0*

To analyse how local demographics impact transmission dynamics, we ran independent simulations using local distribution of demographic characteristics in Birmingham, Leicester, Liverpool, London, Manchester, and York (Supplementary Section S11).

Across all values of *R_0_*, the relative attack rate in non-White ethnic groups was highest in Birmingham (Figure 4 and Supplementary Section S12), where the age gap between individuals from the White ethnic group and other ethnicities was largest, and mean household size was much higher in Asian and Other ethnic groups (Supplementary Figure S18). In Birmingham, Leicester, London and Manchester, the relative attack rate in individuals of Mixed and Black ethnicity was consistently above 1.5 for *R_0_* below 2. In Liverpool and York, cities with the highest proportion of White individuals, the relative attack rate was lower in all ethnic groups. The relative attack rate among individuals of Asian ethnicity was highest in Birmingham and Leicester, both cities where the proportion of individuals of Asian ethnicity is highest, while it was below 1 in York. Age-standardised relative attack rates showed local variations, with lower values for all nonWhite ethnic groups in Liverpool and York compared to other cities (Supplementary Figure S22).

**Figure 4:**
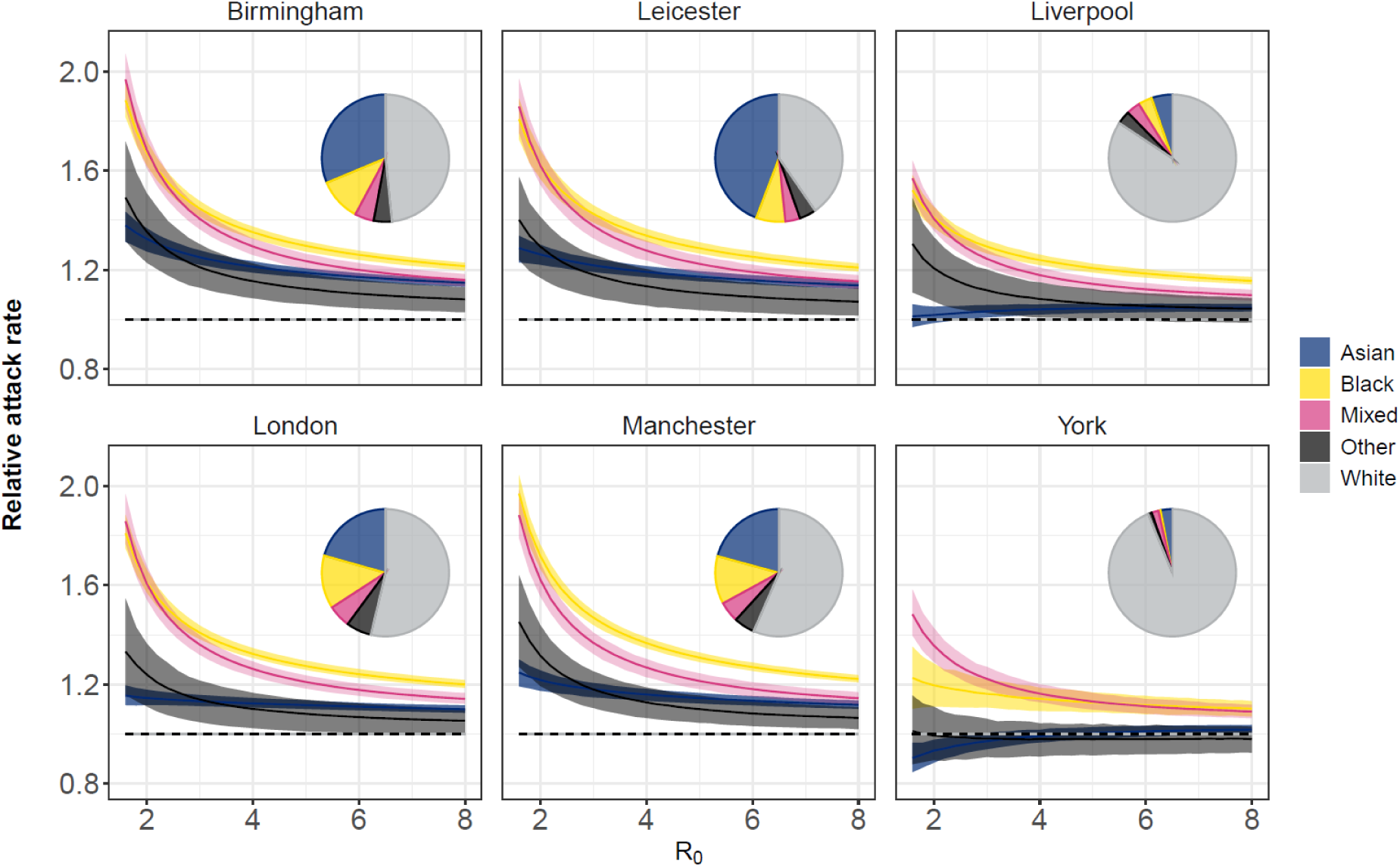
Relative attack rate compared to the White ethnic group for each city and value of R*_0_*. The lines represent the median values, and the ribbons show the 95% Simulation intervals. Inserts show the pie chart of the distribution of the population by ethnicity in each city (from 2021 Census data).

Local demographic distributions led to differences in transmissibility and overall burden (Figure 5). Across all *β*, the lowest *R_0_* was observed in York: *R_0_* was 3.8% [95% SI: -0.6%-7.2%] lower than in England. The highest values of *R_0_* were observed in London and Manchester: 4.5% [95% SI: -0.6%-7.4%] and 5.2% [95% SI: 1.9%-8.7%] higher than in England, respectively. The overall attack rate changed depending on *β*: When *β* was low (with *R_0_* between 1.4 and 1.7 depending on the city), the highest burden was observed in London (19.7% [95% SI: 18.9%-20.4%] of the population infected) and Manchester (20.1% [95% SI: 19.1%-21%] of the population infected). With *β* = 0.039 (*R_0_* between 4.3 and 5 depending on the city), the attack rates in Birmingham, Leicester, London, and Manchester were between 59.9% and 61.0%, while they ranged between 57.9% and 59.4% in England, Liverpool, and York.

**Figure 5:**
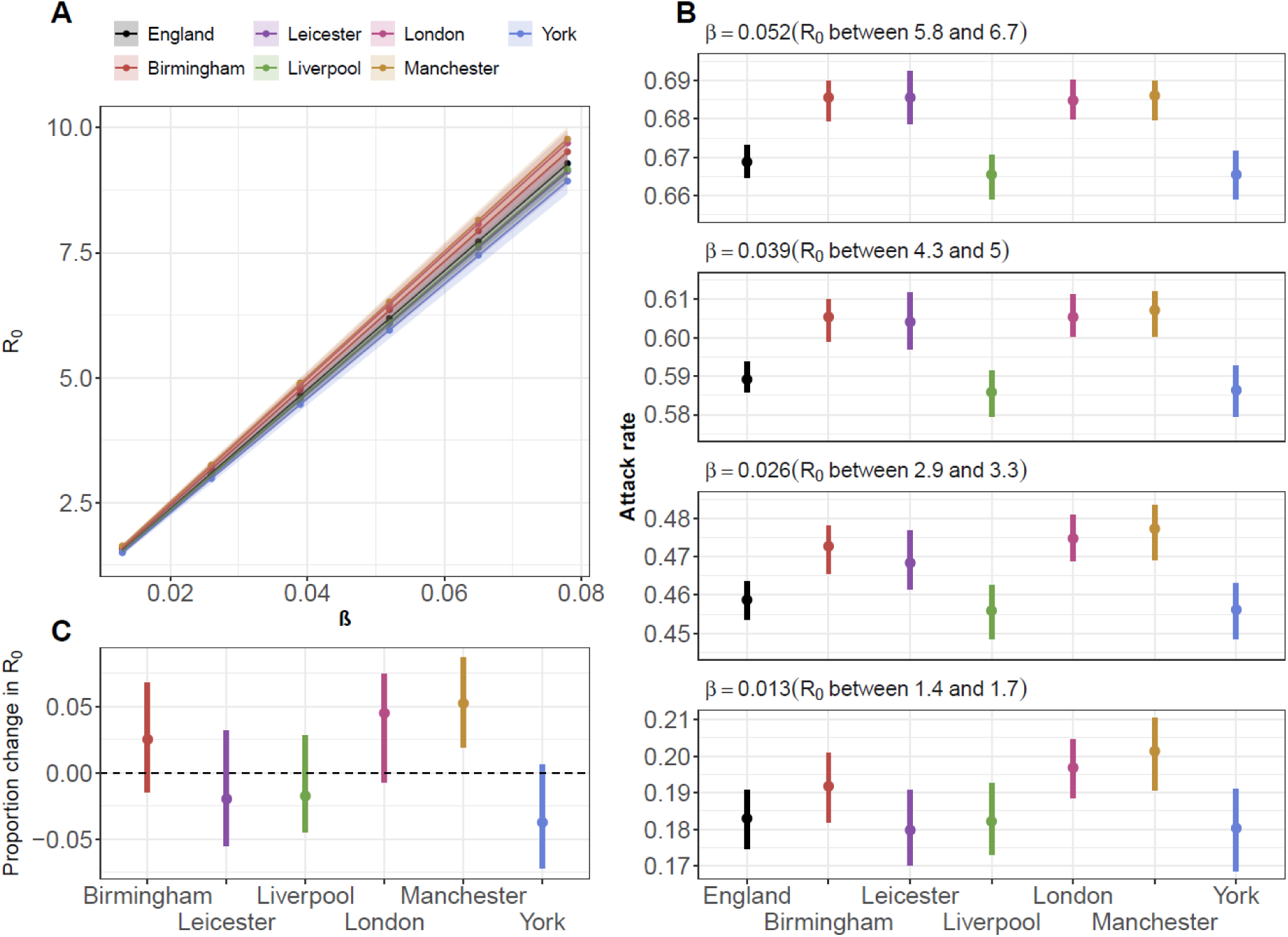
Overall attack rate and R*_0_* by city and. *β*. Top left panel: Value of R*_0_* by *β* for each city. Points show the median value, and ribbons show the 95% Simulation Interval. Bottom left panel: Proportion of change in R*_0_* compared to the value of R*_0_* in England for each city (across all values of *β*). Right panels: overall proportion of the population infected by city for various values of *β*. Points show the median value, and lines show the 95% Simulation Interval.

## Discussion

We found that differences in the distribution of contacts between ethnicities remained after adjusting for the age, employment status, gender, household size and household income of participants. In simulations from an age-, ethnicity- and contact-level stratified model, differences in contact patterns and demographic distributions led to different attack rates between ethnic groups. Demographic factors associated with higher number of contacts among the Reconnect participants were more common in non-White ethnic groups (e.g. younger age groups, larger household size), which led to higher attack rates. The simulated epidemic dynamics were in line with expected trends : high contact groups were at elevated risk of infection, infection rates were higher in younger age groups (17), and individuals of White ethnicity had the lowest rate of infection (1,2).

Previous studies on influenza and COVID-19 incidence highlighted a high incidence in the South Asian ethnic group, while our simulation scenarios showed similar attack rates between the Asian and White groups in various settings. However, direct comparisons are difficult: in the 2011 Census, the ‘South Asian ethnic group’ became the ‘Asian ethnic group’, with the Chinese subgroup moving from Other ethnicity to Asian ethnicity (making up for 8% of Asian ethnicity in 2021) (18). The risk of influenza and COVID-19 infection in the Chinese ethnic group was lower than in other Asian ethnic groups (1,2). The Reconnect study did not report the ethnic subgroup of participants in children (only Asian, Black, Mixed, Other and White groups are available), so we could not stratify our regression analysis by ethnic subgroup. We did not include non-pharmaceutical interventions, control measures, or vaccination in the model, each of which would impact age and ethnic groups in a different way (19,20). We also focused on infections, and therefore did not consider potential differences by age or ethnicity in susceptibility or in propensity for severe outcomes, which would change the observed burden of disease.

Local differences in distribution of age and ethnicity by city changed the level of inequalities in simulated epidemics. The relative attack rate compared to the White ethnic group reached 2 in Black and Mixed groups in Birmingham for low values of *R_0_*, but remained below 1.6 in all groups in York. Heterogeneity between age, ethnicity, and contact group distributions between cities also impacted *R_0_*: when the transmissibility rate was held constant, *R_0_* was 8.7% [95% SI: 6.1% - 11.3%] higher in Manchester than in York, and translated into an absolute difference in overall incidence of 2 to 3% between cities. This would slightly affect the level of control measure adherence and vaccine coverage needed to mitigate outbreak risks depending on the area (21). In all simulations, the relative attack rate between ethnicities varied with *R_0_*, for instance, at a national level, the attack rate in individuals of Asian ethnicity was close to that of individuals of White ethnicity for *R_0_* below 3, but similar to individuals of Mixed ethnicity when *R_0_* was above 6. Taken together, our findings indicate that the inequalities in infection incidence due to demography, social mixing, and contact distribution alone are expected to vary across settings and pathogens.

There were three sources of heterogeneity in the model: the distribution of demographic characteristics, the remaining differences in observed contact rates between ethnic groups after adjusting for demographic variables, and the mixing matrix between ethnicities. To assess the share of heterogeneity due to the residual differences between ethnic groups estimated by the regression model, we used the national level demographic characteristics of the population and used homogeneous mixing between ethnicities. This resulted is substantially more homogeneous transmission dynamics, which shows that factors not included in the regression analysis should have a limited impact on simulated transmission dynamics. Adding factors such as the type of employment and measures of socioeconomic background in the regression analysis could have further reduced the remaining differences in contact distribution between ethnicities after adjusting for demographic variables (22).

The regression analysis found that household size was associated with an increased number of contacts, but individuals from households of four and more than four individuals had a similar number of contacts. This observation mirrors previous studies highlighting a “plateaued” effect of household size on influenza transmission (23). Other studies have shown the importance of household structure, instead of just household size, on transmission risk, since age impacts the risk of exposure to pathogens (24). Variables on household composition were not integrated in the regression analysis, and age group mixing may also differ in multi-generational households. We found that the number of contacts increased with annual household income, despite COVID-19 and influenza epidemics generally showing higher disease burden in people living in more deprived areas (25,26). This discrepancy may be due to factors not included in this analysis, for example vaccine coverage, which varies across socioeconomic groups (27), or opportunities to isolate and follow non-pharmaceutical interventions (28).

Implementing a compartmental model stratified by age, ethnicity, and contact group required computing the mixing matrix between each stratum. To do so, we combined separate mixing matrices by age and by ethnicity from Reconnect, and the number of contacts per contact group. We assumed that the age-specific mixing was the same for all ethnicities and contact groups, and that ethnicity-specific mixing was the same for all age and contact groups. In particular, mixing patterns in school-age children may differ from the rest of the population and rely on geographical locations and school catchment areas (29). The different contact groups may also not all behave in the same way: in our simulation, social contacts from high contact groups were distributed following the distribution in the reconnect survey, but high contact groups may follow a different assortativity of mixing. As highlighted by the different scenario and previous analyses (13,30), ethnic assortativity is one of the drivers of heterogeneous transmission dynamics. We also assumed that the per capita mixing matrices were the same in all cities where we ran the model.

This study describes factors influencing the distribution of contacts between ethnicities in a large representative study in England. We show how heterogeneity in demography, mixing patterns and number of contacts translate into inequalities in infection incidence between ethnic groups, and potentially different local epidemic dynamics. The main extension of these results will be to fit the transmission model to case data to better understand age-specific mixing patterns between ethnicities, integrate the impact of inequalities in vaccine coverage and control measures, and inequalities in severity of outcomes.

## Data Availability

The code, along with instructions to reproduce the contact matrices, regression analysis compartmental models and simulations can be found at https://github.com/alxsrobert/connect_overdispersion. The full individual-level Reconnect contact dataset cannot be shared publicly for the protection of participants privacy. To make the study as reproducible as possible, we generated a simulated contact dataset and included it in the Github repository, so readers can generate model fits and simulations.

## Acknowledgements

We would like to thank Professor Rohini Mathur for useful discussions and feedback on the article.

## Data sharing

The code, along with instructions to reproduce the contact matrices, regression analysis compartmental models and simulations can be found at https://github.com/alxsrobert/connect_overdispersion. The full individual-level Reconnect contact dataset cannot be shared publicly for the protection of participants’ privacy. To make the study as reproducible as possible, we generated a simulated contact dataset and included it in the Github repository, so readers can generate model fits and simulations.

