## supplementary material for "Ethnic inequalities in respiratory virus epidemics in England: a mathematical modelling study"

#### Supplementary Section S1. Project workflow

**Figure S1:Schematics showing the research questions, data sources, and methods used in the analysis.**

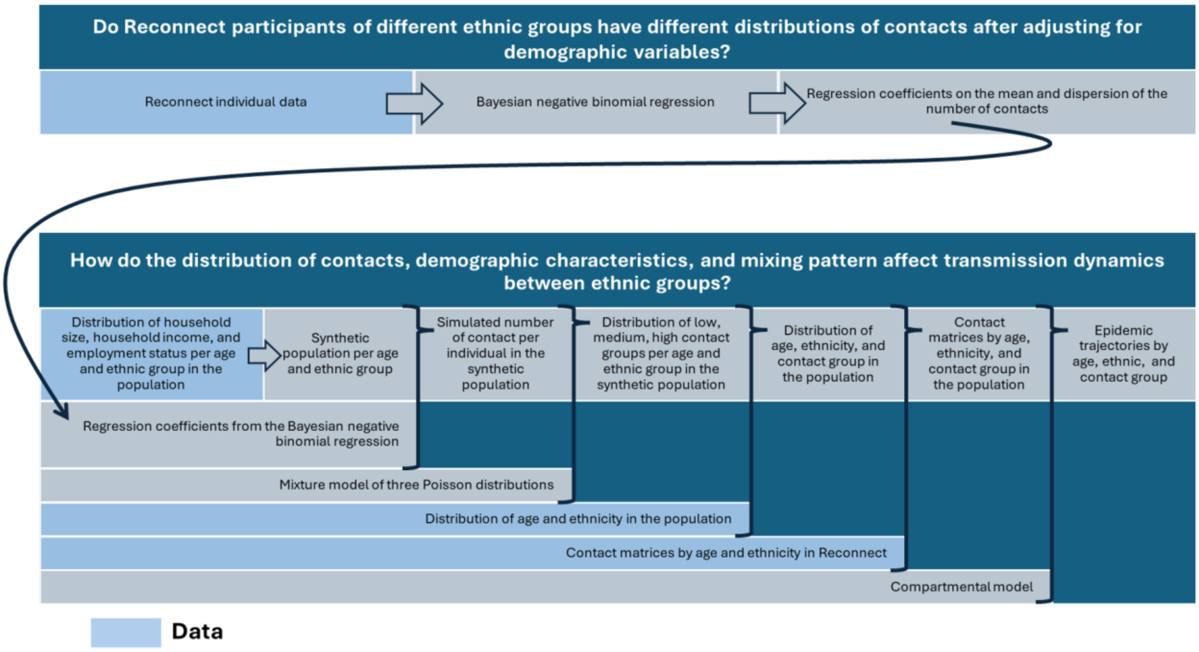

#### Supplementary Section S2. Distribution of variables in Reconnect

In Reconnect, participants recorded their own characteristics as well as their contacts' over a 24-hour period. Two types of contact were recorded in Reconnect: individual contacts, for which the contacts' age group and ethnicity were collected, and large group contacts, for which only the contacts' broad age groups were collected (under 18, 18-64, 65 and over), with no information on the contacts' ethnicity. In total, 50,665 individual and 75,006 large group contacts from 13,238 participants were collected. The variables used for each participant were their age group (stratified in 11 groups: 0-4; 5-9; 10-14; 15-17; 18-24; 25-29; 30-39; 40-49; 50-59; 60-69; 70+), household size (1, 2, 3, 4, More than 4), ethnic group (Asian, Black, Mixed, Other, White), urban/rural status, annual household income (under £20,000; £20,000-£39,999, £40,000-59,999, £60,000-£99,999; £100,000 and over), gender (Male, Female, Other), whether contact data was collected on weekdays or weekends, and employment status (Employed, Unemployed, Retired, Looking after home or family, Long-term sick or disabled, Students, Other). All full- or part-time, employed or self-employed individuals were classified as "Employed". Adult participants reported their ethnic subgroup (White English / Welsh / Scottish / Northern Irish / British, White Gipsy or Irish Traveller, White Irish, White Other, Bangladeshi, Chinese, Indian, Pakistani, Other Asian background, African, Caribbean, Other Black/African/Caribbean background, White and Asian, White and Black African, White and Black Caribbean, Other mixed / multiple ethnic background, Latin American, Arab, Other ethnic background). Children only reported their ethnic group (Asian, Black, Mixed, Other, White).

The employment status and household income of all participants below 18 were classified as "Child (Not Applicable)" in Reconnect. Urban/rural status was derived from the first half of the postcode (see Supplementary materials of Goodfellow et al (1)). Due to missingness or an unmatched postcode, the urban/rural status could not be computed for 736 participants (5.6% of all entries), who were therefore removed from the dataset. We also removed 18 further participants, for which the age group, gender, household size or income, or employment status was not known.

Figure S2 presents some of the demographic characteristics of the remaining 12,484 participants, the dataset used in this study. In Reconnect, 92.8% of participants from rural areas identified their ethnic group as White, in contrast with 77.7% of individuals in urban areas (Supplementary Figure S3), in line with the distribution of ethnicity in urban and rural areas in England. We created a composite variable covariate merging ethnic group and urban/rural status, with six levels: Asian and Urban (7.6% of all individuals in the Reconnect dataset), Black and Urban (9.0%), Mixed and Urban (2.3%), Other and Urban (0.5%) White and Urban (67.7%), non-White and Rural (0.9%), White and Rural (11.9%).

**Figure S2 : Distribution of demographic variables among participants of the Reconnect survey. The fourth row refers to the distribution of household size.**

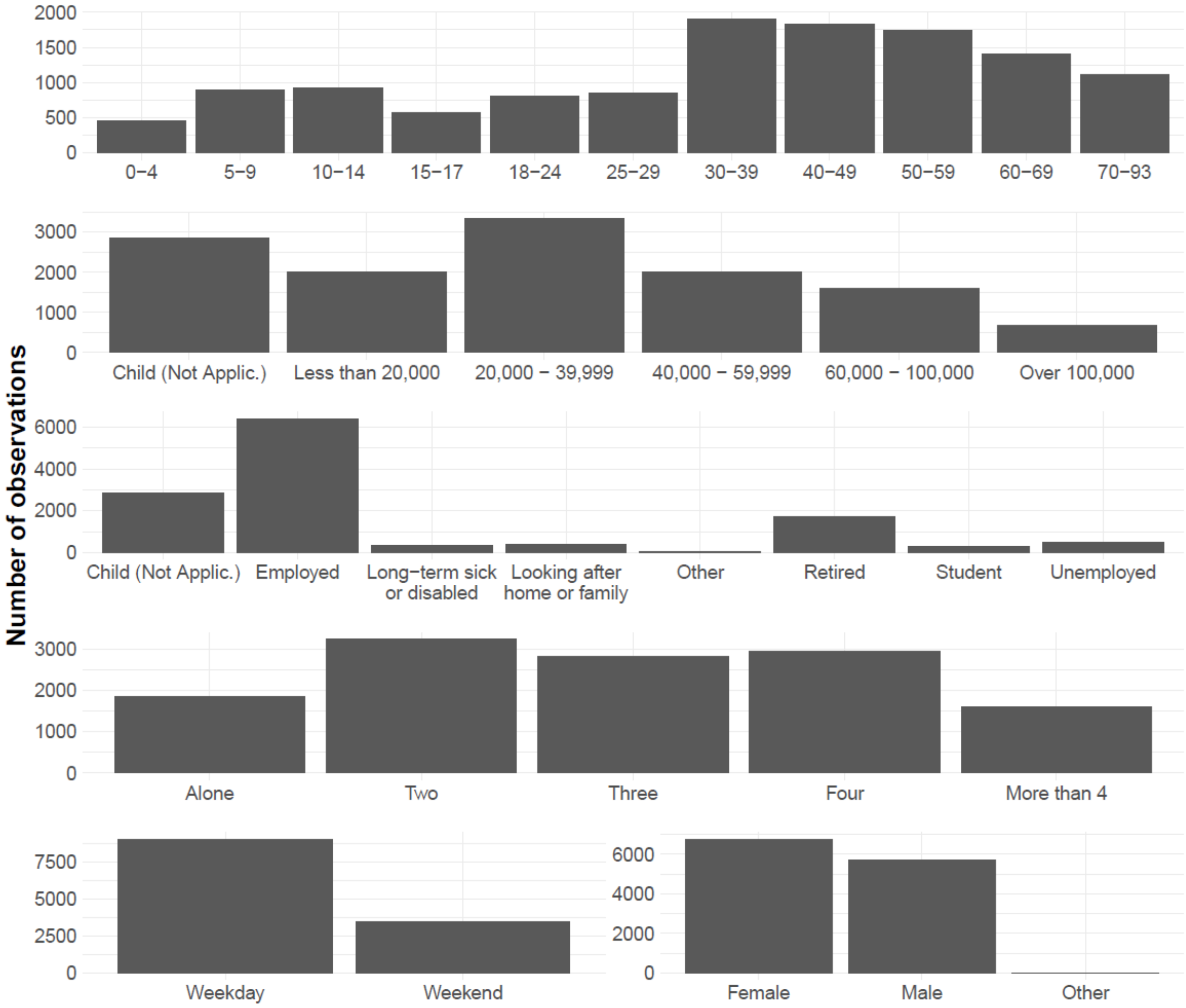


**Figure S3: Distribution of ethnicity in the Reconnect contact survey, by urban and rural status. The label corresponds to the percentage in each location.**

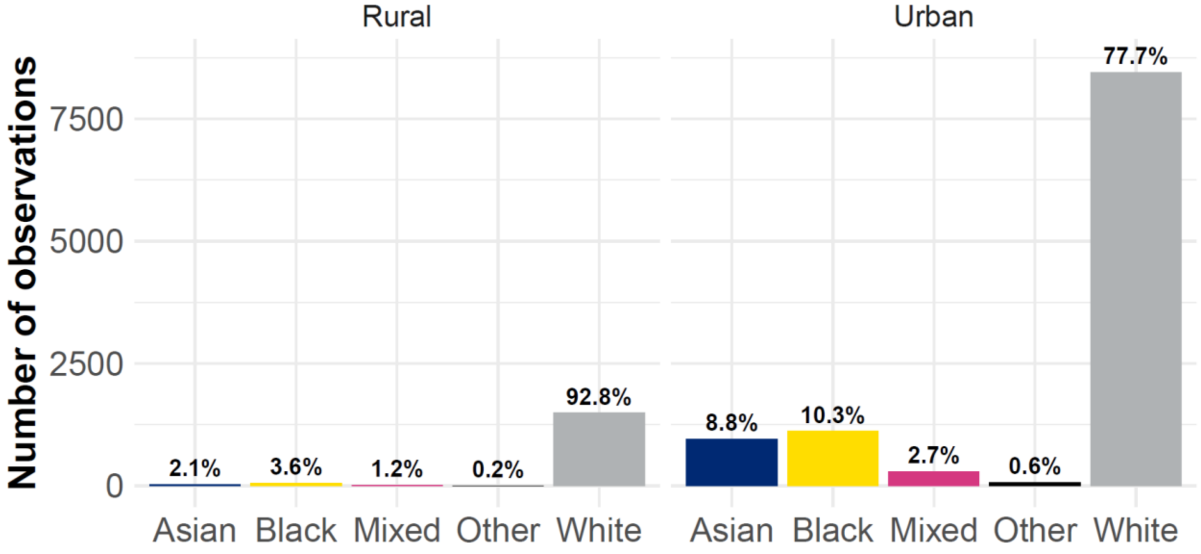

#### Supplementary Section S3. Regression model: Description and equations

In Reconnect, the employment status and annual household income of all participants below 18 was classified as "Child - Not applicable" (see Supplementary Section S2). Including these levels in the regression model introduced collinearity with children's age levels. To mitigate this, we excluded "Child - Not applicable" from the levels of household income and employment status. Age group coefficients for participants below 18 therefore reflect both age effects, and the impact of being outside the reference level of employment status and household income.

When estimating the association between the mean number of contacts and the urban/rural - ethnicity status, we adjusted for various demographic variables collected in Reconnect: participant's age group (reference level: 18-24), employment status (reference level: Employed), annual household income (reference level: £20,000-£39,999), household size (reference level: 1), and gender (reference level: Female), and whether the survey day was a weekday (reference: yes). All model parameters were set up with a uniform prior distribution. We also estimated the association between the dispersion parameter and each level of urban/rural ethnicity status.

The number of contacts for each individual $i$, $y_i$, is modeled as: $y_i \sim NegBin(\mu_i, \phi_i)$, with $\mu_i$ the mean number of contact, and $\phi_i$ the dispersion parameter. The equations of the Negative Binomial regression model are:

$$\begin{aligned} log(\mu_i) = \beta_0 &+ \beta_1 \cdot \{AsianUrban\}_i + \beta_2 \cdot \{BlackUrban\}_i + \beta_3 \cdot \{MixedUrban\}_i \\ &+ \beta_4 \cdot \{OtherUrban\}_i + \beta_5 \cdot \{WhiteRural\}_i + \beta_6 \cdot \{NonWhiteRural\}_i \\ &+ \textstyle\sum_{j=1}^{11} \gamma_j \cdot \{AgeGroup\}_{ij} + \sum_{k=1}^{6} \delta_k \cdot \{EmployStatus\}_{ik} + \sum_{l=1}^{4} \theta_l \cdot \{Income\}_{il} \\ &+ \textstyle\sum_{m=1}^{4} \eta_m \cdot \{HHSize\}_{im} + \zeta_1 \cdot \{Male\}_i + \zeta_2 \cdot \{OtherGender\}_i + x_i \cdot \{Weekend\}_i \end{aligned}$$

$$\begin{aligned} log(\phi_i) = \alpha_0 &+ \alpha_1 \cdot \{AsianUrban\}_i + \alpha_2 \cdot \{BlackUrban\}_i + \alpha_3 \cdot \{MixedUrban\}_i \\ &+ \alpha_4 \cdot \{OtherUrban\}_i + \alpha_5 \cdot \{WhiteRural\}_i + \alpha_6 \cdot \{NonWhiteRural\}_i \end{aligned}$$

All $\alpha$, $\beta$, $\gamma$, $\delta$, $\theta$, $\eta$, $\zeta$ are coefficients of the model, estimated in the inference process. The subscripts correspond to categorical variables: $j$ (age group), $k$ (employment status), $l$ (income category), $m$ (household size).

#### Supplementary Section S4. Computing contact groups per age and ethnicity stratum

To capture the dispersion of contact distribution in a way that can be easily embedded in a compartmental epidemic model, individuals in each age and ethnicity stratum were further stratified by contact group. Individuals belonging to the same stratum of a compartmental model have the same number of contacts and mixing patterns, so the stratification by contact groups allowed for different behaviours within the same stratum. To create the contact groups, we used synthetic populations with 500 individuals for each age-ethnicity stratum. For each level of age and ethnicity, individuals in the synthetic population were assigned a gender, household size (using the distribution by age and ethnicity in the area of interest from the 2021 Census), employment status (using the distribution by age, gender and ethnicity in the area of interest from the 2021 Census), annual income category (using the distribution by ethnicity in England from the Family Resources Survey). We used the regression outputs to simulate the number of contacts per individual. In each level of age and ethnicity, we then fitted a mixture model to the simulated distribution of contacts in the synthetic population. The mixture model comprised three Poisson distributions. In each level of age and ethnicity, the parameters of the mixture model corresponded to the average number of contacts, and the proportion of the population belonging to the low, medium, and high contact groups. Supplementary Figures S4 to S9 show the density plots of the number of contacts per age group and ethnicity in the synthetic populations and mixture models. We implemented a sensitivity analysis using four Poisson distributions, which did not change the relative attack rates between ethnicities in the simulations (Supplementary Section S10).

**Figure S4: Density plots of the number of contacts among 0-4 and 5-9 year old individuals, stratified by ethnicity.** The blue density shows the distribution of contacts in the synthetic population, simulated from the distribution of demographic variables in England, and the outputs from the regression analysis. The yellow, orange, and red densities show the distribution of contacts in the three Poisson distributions of the mixture model fitted separately for each age group and ethnicity. The brown density shows the weighted mixture distribution.

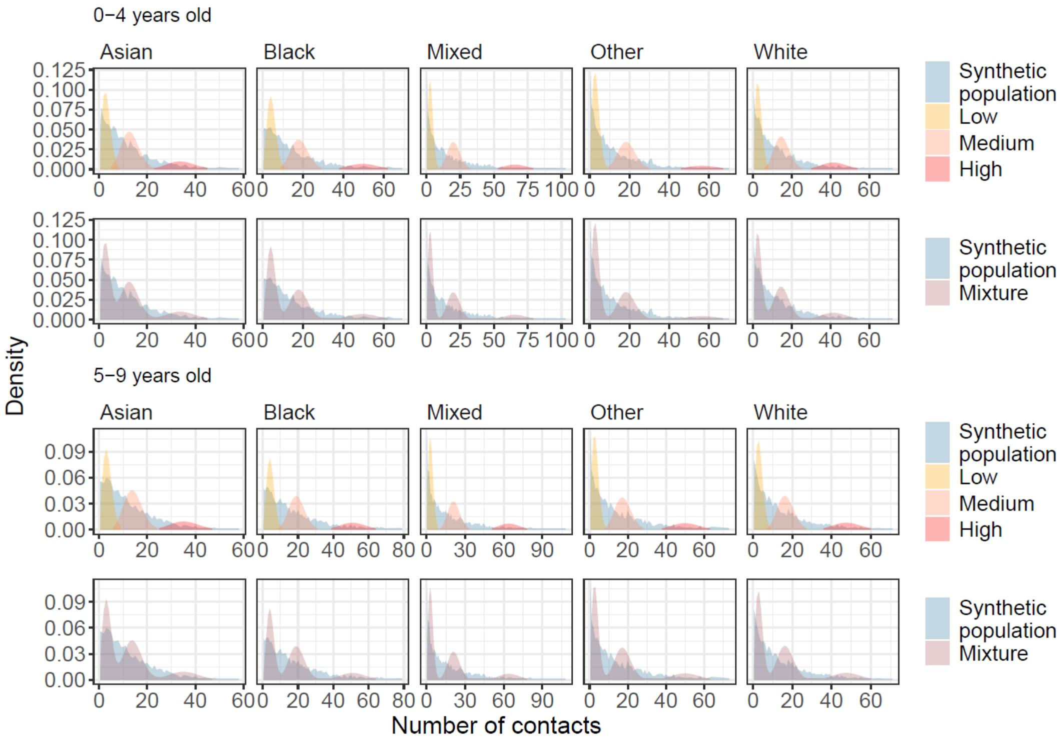

**Figure S5: Density plots of the number of contacts among 10-14 and 15-17 year old individuals, stratified by ethnicity.** The blue density shows the distribution of contacts in the synthetic population, simulated from the distribution of demographic variables in England, and the outputs from the regression analysis. The yellow, orange, and red densities show the distribution of contacts in the three Poisson distributions of the mixture model fitted separately for each age group and ethnicity. The brown density shows the weighted mixture distribution.

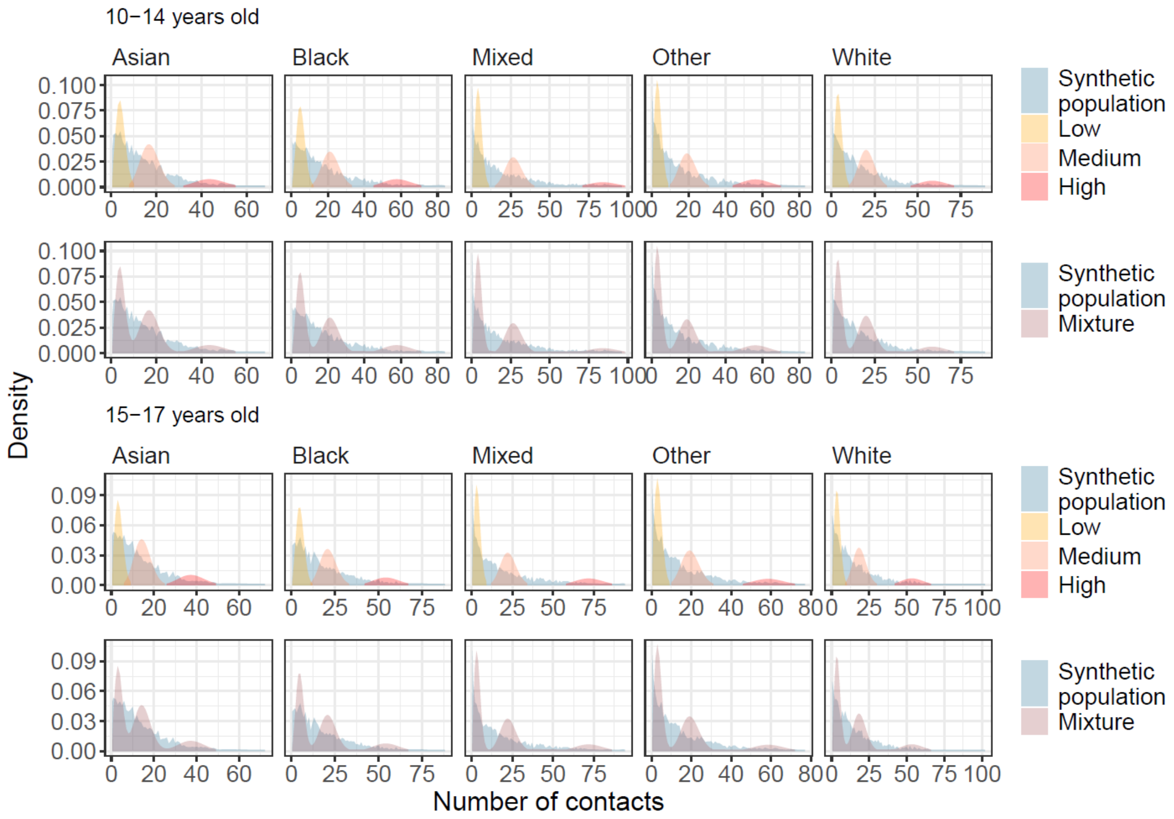

**Figure S6: Density plots of the number of contacts among 18-24 and 25-29 year old individuals, stratified by ethnicity.** The blue density shows the distribution of contacts in the synthetic population, simulated from the distribution of demographic variables in England, and the outputs from the regression analysis. The yellow, orange, and red densities show the distribution of contacts in the three Poisson distributions of the mixture model fitted separately for each age group and ethnicity. The brown density shows the weighted mixture distribution.

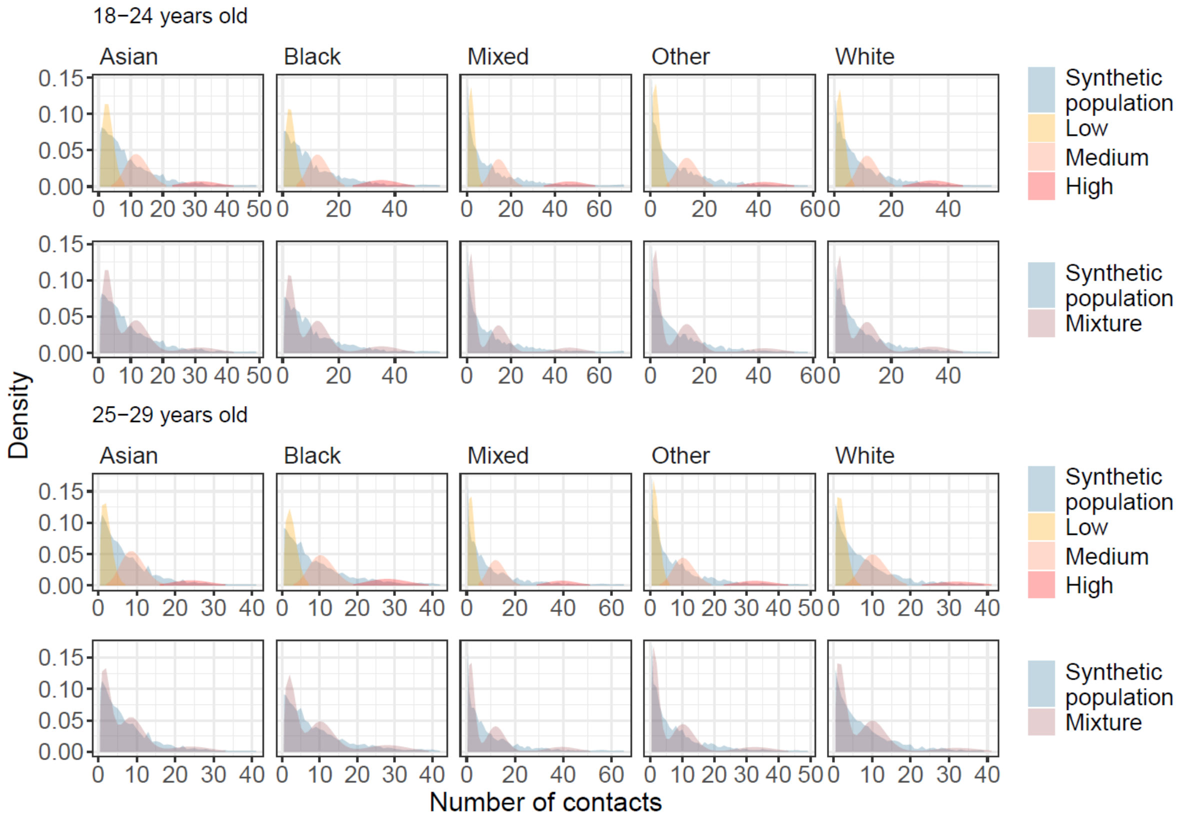

**Figure S7: Density plots of the number of contacts among 30-39 and 40-49 year old individuals, stratified by ethnicity.** The blue density shows the distribution of contacts in the synthetic population, simulated from the distribution of demographic variables in England, and the outputs from the regression analysis. The yellow, orange, and red densities show the distribution of contacts in the three Poisson distributions of the mixture model fitted separately for each age group and ethnicity. The brown density shows the weighted mixture distribution.

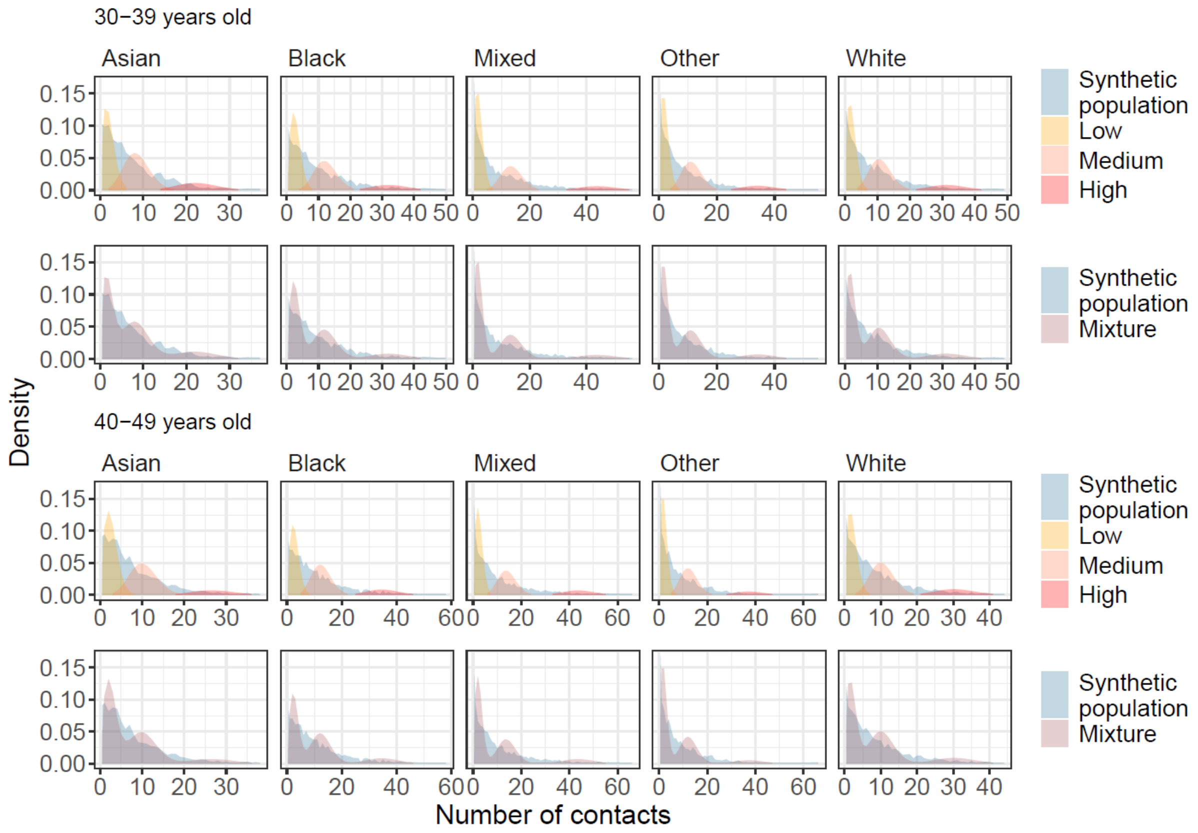

**Figure S8: Density plots of the number of contacts among 50-59 and 60-69 year old individuals, stratified by ethnicity.** The blue density shows the distribution of contacts in the synthetic population, simulated from the distribution of demographic variables in England, and the outputs from the regression analysis. The yellow, orange, and red densities show the distribution of contacts in the three Poisson distributions of the mixture model fitted separately for each age group and ethnicity. The brown density shows the weighted mixture distribution.

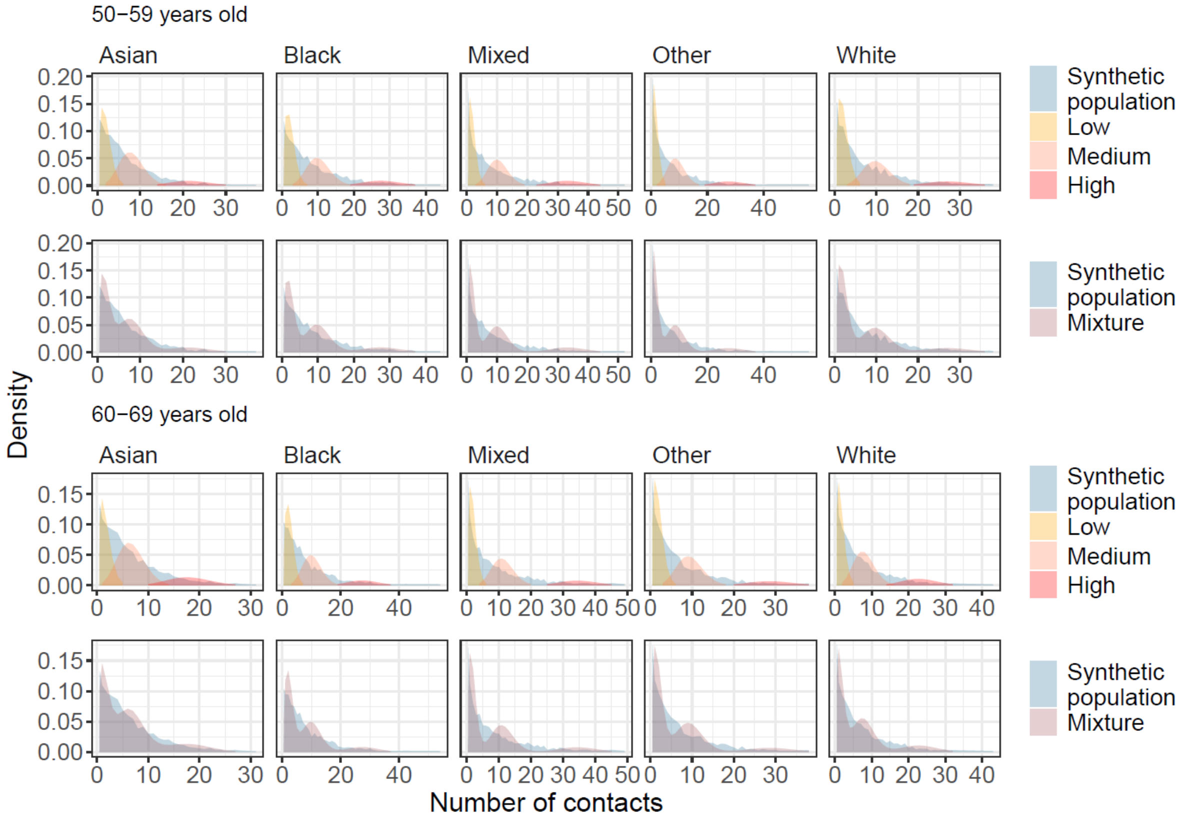

**Figure S9: Density plots of the number of contacts among 70-93 year old individuals, stratified by ethnicity.** The blue density shows the distribution of contacts in the synthetic population, simulated from the distribution of demographic variables in England, and the outputs from the regression analysis. The yellow, orange, and red densities show the distribution of contacts in the three Poisson distributions of the mixture model fitted separately for each age group and ethnicity. The brown density shows the weighted mixture distribution.

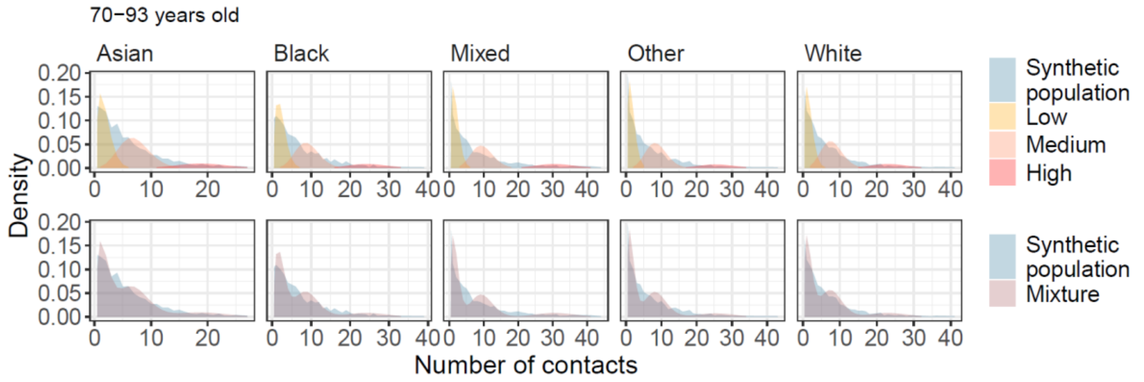


### Supplementary Section S5. Computing the per capita matrix stratified by age group, ethnicity, and contact group

We built an SEIR simulation model, stratified by age group, ethnicity, and contact group (i.e. low, medium, and high number of contacts). The proportion of the population in the low / medium / high contact group and the number of contacts associated with each group varies by age group and ethnicity. To build this model, we constructed the per capita contact matrix stratified by age group, ethnic group and number of contacts, built from the age-stratified and ethnicity-stratified contact matrices from Reconnect, and the distribution of contacts by age and ethnicity in the synthetic population. In Reconnect, the ethnicity and age groups were recorded for all participants, but not for all contacts: contacts classified as large group contacts only have their broad age groups recorded, but not ethnicity. Therefore, we did not directly construct a joint age-ethnicity contact matrix from the Reconnect data. Instead we used the individual contacts (as opposed to large group contacts) to compute the distribution of contacts between ethnic groups (i.e. the proportion of all contacts between each ethnic group).

We assumed that the age, ethnic, and contact stratification of contacts were independent. For instance, the per capita contact rate between any pair of ethnic groups was the same across all age and contact groups (e.g. all individuals of White ethnicity may have 0.0001 contacts per person of Black ethnicity, regardless of their age and contact group).

Combining all stratifications was a complex process, described in Figure S10.

**Figure S10: Workflow describing how the per capita matrix was computed**

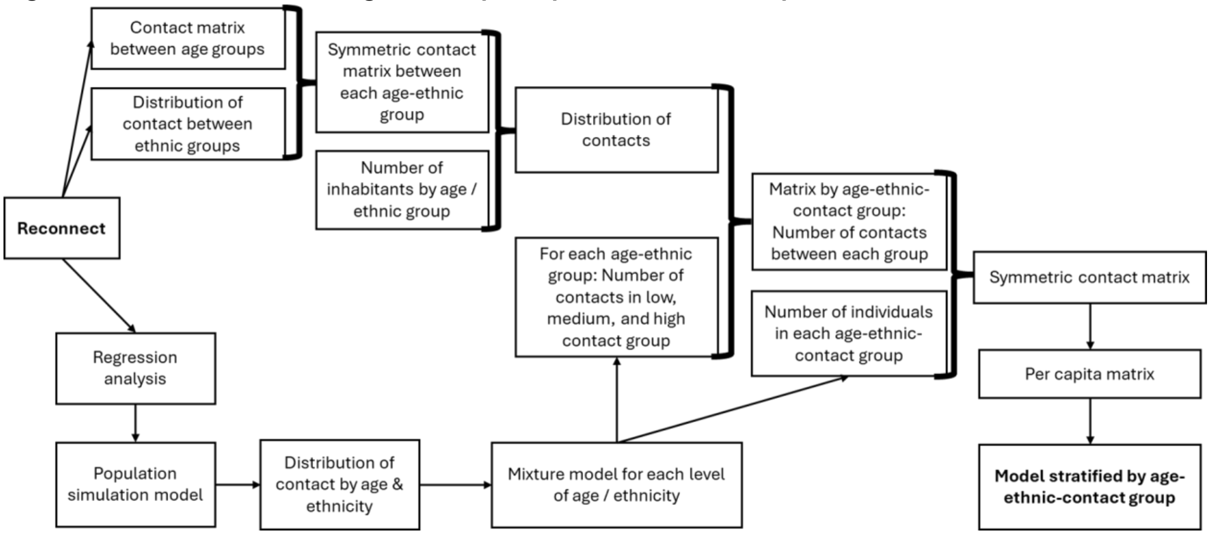


Here we present the detailed steps of the computation, with parameters:

- Age groups: $i, j \in \{1, \ldots, I\}$
- Ethnic groups: $k, l \in \{1, \ldots K\}$
- Contact groups: $g, h \in \{low, med, high\}$.
- Total population in Reconnect: $N^{rec}$
- Total population in the population of interest: $N^{2024}$
- Population of age $i$: ${N_i}^{2024}$
- Population of age $i$ and ethnic group $k$: ${N_{i,k}}^{2024}$
- Average number of contacts from age $i$ to age $j$ in Reconnect: ${M_{ij}}^{rec}$
- Average number of contacts from age $i$ to age $j$ in the population of interest: ${M_{ij}}^{2024}$
- Average frequency of contacts from age $i$ to age $j$ in Reconnect (i.e. per capita contact matrix): ${P_{ij}}^{rec}$
- Average frequency of contacts from age $i$ to age $j$ in the population of interest (i.e. per capita contact matrix): ${P_{ij}}^{2024}$

1. **Construct the symmetric contact matrix stratified by age and ethnicity** (top left side of Supplementary Figure S10)
   A. **Project the age and ethnicity per-capita matrix** (Arregui et al(2)): ${P_{ij}}^{2024} = {P_{ij}}^{rec} * \frac{N^{rec}}{N^{2024}}$, ${P_{kl}}^{2024} = {P_{kl}}^{rec} * \frac{N^{rec}}{N^{2024}}$
   B. **Compute the average number of contacts per participant:** ${M_{ij}}^{2024} = {P_{ij}}^{2024} * {N_j}^{2024}$
   C. **Distribute age-stratified contacts between ethnicities**
      i. Compute $T_{ij}$ the total number of contacts from all participants aged $i$ to contacts aged $j$: $T_{ij} = {M_{ij}}^{2024} * {N_i}^{2024}$.
      ii. Compute the distribution of contacts by ethnicity given the distribution of ethnicity in age groups $i$ and $j$: ${C_{kl}}^{ij} = \frac{{P_{kl}}^{2024} * N_{ik} * N_{jl}}{\sum_{k'=1}^{K} \sum_{l'=1}^{K} {P_{k'l'}}^{2024} * N_{ik'} * N_{jl'}}$.

iii. Compute the total number of contacts between each ethnic and age group: $T_{ik,jl} = T_{ij} * C_{kl}{}^{ij}$

iv. Compute the average number of contacts from participants aged $i$ of ethnicity $k$ to contacts aged $j$ of ethnicity $l$: $M_{ik,jl} = T_{ik,jl}/N_{ik}$

2. **Create contact groups** (bottom side of Supplementary Figure S10). For each stratum $(i, k)$ (age-ethnicity):
   A. Generate $S = 500$ individuals per stratum $(i, k)$. For each individual $s$ in stratum $(i, k)$, draw: $sex_s \sim Bernoulli(0.5)$, $household_s \sim Census(age = i, ethnicity = k)$, $employment_s \sim Census(age = i, ethnicity = k, gender = gender_s)$, $income \sim survey(ethnicity = k)$.
   B. Draw the number of contacts ($Y_{n=1…2500}$) given the parameters from the regression outputs (using five draws) and the characteristics of each individual (2500 contact counts per stratum).
   C. Fit a mixture of three Poisson distributions to the 2500 contact counts in each stratum: $\lambda_{ikg}$ is the mean number of contacts in contact group $g$ (low/medium/high) for stratum $(i, k)$, $\pi_{ikg}$ is the proportion of individuals in contact group $g$ for the stratum $(i, k)$.
3. **Compute the age-ethnicity-contact group matrix** (right-hand side of Supplementary Figure S10):
   **A. Compute the distribution of contacts in each age-ethnicity stratum**: $q_{ik,jl} = \frac{M_{ik,jl}}{\sum_{j'=1}^{I} \sum_{l'=1}^{K} M_{ik,j'l'}}$. So $q_{ik,jl}$ is the proportion of contacts of an individual in $(i, k)$ that go to individuals in $(j, l)$.
   **B. Compute the number of contact between each age, ethnicity, and contact stratum**
      **i. Compute the average number of contacts between the participant's age-ethnicity-contact group and contact's age-ethnicity group:** Individuals in $(i, k, g)$ have $\lambda_{ikg}$ contacts on average. The number of contacts from $(i, k, g)$ to $(j, l)$ is computed as: $M_{ikg,jl} = \lambda_{ikg} * q_{ikjl}$.
      **ii. Compute the weighted distribution of contacts over contact groups**: Within contact's age-ethnicity stratum $(j, l)$, we use the proportion of individuals in each contact group, and the mean number of contacts per group: $w_{jlh} = \frac{\pi_{jlh}\lambda_{jlh}}{\sum_{h'=1}^{3} \pi_{jlh'}\lambda_{jlh'}}$. This accounts for the lower probability of "reaching" low-contact individuals (proportionate mixing).
      **iii. Compute the average number of contacts stratified by age-ethnicity-contact group:** $M_{ikg,jlh} = M_{ikg,jl} * w_{jlh}$.
   **C. Make the age-ethnicity-contact group matrix symmetrical:** $M_{ikg,jlh}{}^{standard} = (M_{ikg,jlh} * N_{ikg} + M_{jlh,ikg} * N_{jlh}) * \frac{1}{N_{ikg}}$.
   **D. Compute the per capita matrix:** $P_{ikg,jlh} = \frac{M_{ikg,jlh}{}^{standard}}{N_{jlh}}$

Combining the mixing matrices, population structure, and number of contacts led to differences between the final contact matrix (calculated in step 3) and the number of contacts in the mixture model (calculated in step 2). This is due to differences between the age and ethnicity mixing matrices from Reconnect (calculated in step 1) and the contact distributions in the mixture model. The comparison between the number of contacts per group in the mixture model and in the final per capita matrix is shown in Figure S11, and highlights that the differences were minimal across age and ethnic groups.

**Figure S11: Comparison of the mean number of contacts per age group, ethnicity, and contact group in the mixture distribution and in the per capita contact matrix used in the SEIR simulation model.** The per capita contact matrix stratified by age group, ethnicity, and contact group are computed from the age and ethnicity per capita contact matrices from the Reconnect study, and the number of contacts by contact group. Changes in the mean number of contacts are introduced when distributing the number of contacts from the contact groups by age and ethnicity of contacts (Step 3/A and 3/B), and making the resulting per capita matrix symmetrical (Step 3/C).

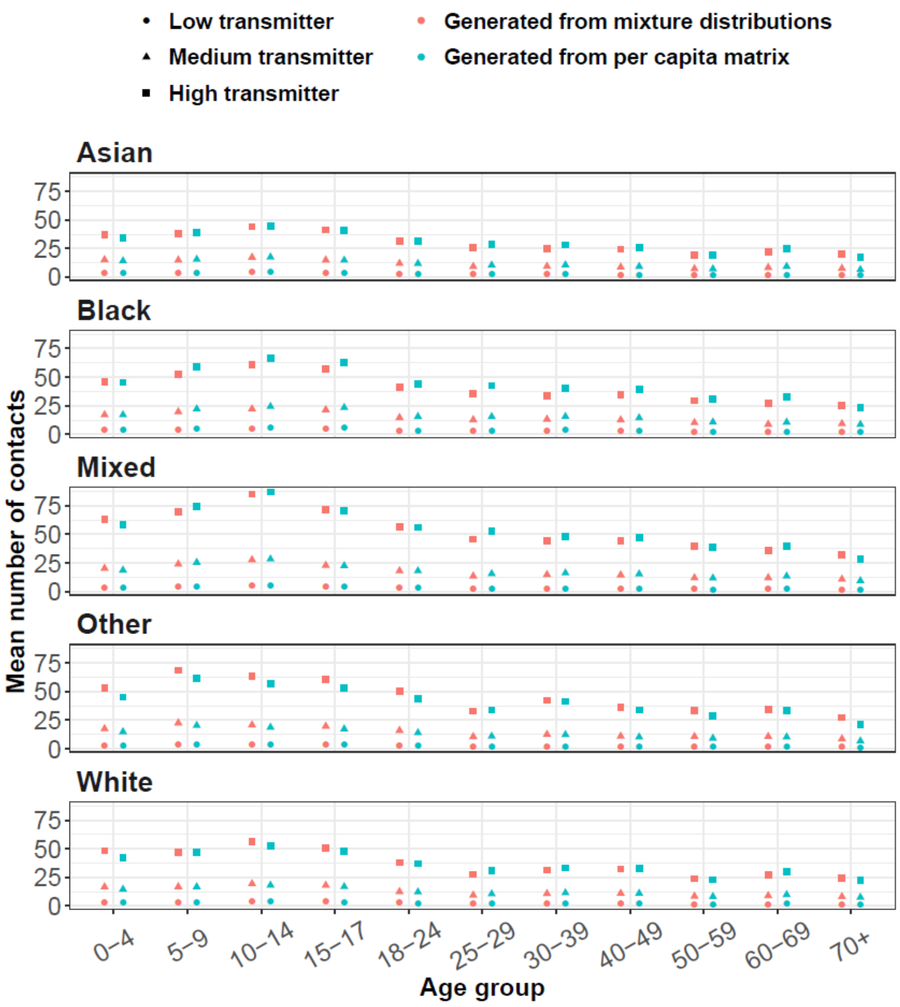


#### Supplementary Section S6. Regression outputs: Parameter estimates

Supplementary Table S1 presents the mean and 95% Credible Interval of all coefficients in the full regression model.

**Table S1: Parameter estimates of the regression model.**

| Covariate | Term | Value [95% CI] |
|---|---|---|
| Intercept | (Intercept) | 7.24 [6.51 - 8.08] |
| Ethnicity and urban / rural | Asian and Urban | 0.85 [0.79 - 0.91] |
| | Black and Urban | 1.18 [1.11 - 1.26] |
| | Mixed and Urban | 1.31 [1.14 - 1.52] |
| | Other and Urban | 1.03 [0.78 - 1.39] |
| | Nonwhite and Rural | 1.24 [1.02 - 1.53] |
| | White and Rural | 0.95 [0.89 - 1.01] |
| Age | 0-4 years old | 1.33 [1.16 - 1.52] |
| | 5-9 years old | 1.44 [1.28 - 1.61] |
| | 10-14 years old | 1.63 [1.46 - 1.83] |
| | 15-17 years old | 1.56 [1.37 - 1.77] |
| | 25-29 years old | 0.9 [0.81 - 1] |
| | 30-39 years old | 0.98 [0.89 - 1.08] |
| | 40-49 years old | 1.01 [0.92 - 1.12] |
| | 50-59 years old | 0.84 [0.76 - 0.93] |
| | 60-69 years old | 0.89 [0.79 - 1] |
| | 70+ years old | 0.92 [0.8 - 1.06] |
| Income | Less than £20,000 per year | 0.95 [0.89 - 1.02] |

| | | |
|---|---|---|
| | £40,000-59,999 per year | 1.17 [1.1 - 1.25] |
| | £60,000-99,999 per year | 1.1 [1.03 - 1.17] |
| | Over £100,000 per year | 1.34 [1.22 - 1.48] |
| Employment status | Other | 0.77 [0.54 - 1.12] |
| | Looking after home or family | 0.56 [0.49 - 0.63] |
| | Retired | 0.76 [0.69 - 0.84] |
| | Unemployed | 0.7 [0.63 - 0.78] |
| | Student | 1.36 [1.19 - 1.56] |
| | Long-term sick or disabled | 0.72 [0.63 - 0.83] |
| Household size | Two | 1.28 [1.2 - 1.38] |
| | Three | 1.32 [1.22 - 1.42] |
| | Four | 1.56 [1.44 - 1.69] |
| | More than four | 1.49 [1.37 - 1.63] |
| Day of the week | Weekend | 0.84 [0.8 - 0.88] |
| Gender | Male | 0.84 [0.81 - 0.88] |
| | Other | 2.88 [1.6 - 5.86] |
| Dispersion Parameter | (Intercept) | 0.87 [0.84 - 0.89] |
| | Asian and Urban | 1.37 [1.24 - 1.51] |
| | Black and Urban | 1.26 [1.15 - 1.38] |
| | Mixed and Urban | 0.81 [0.69 - 0.94] |

| | | |
|---|---|---|
| | Other and Urban | 0.94 [0.65 - 1.33] |
| | Nonwhite and Rural | 1.1 [0.85 - 1.42] |
| | White and Rural | 1.04 [0.96 - 1.13] |

Supplementary Figure S12 shows the values of the estimated coefficient in the full regression model (presented in the Main text), and in a regression model without adjustment on demographic variables (i.e. the composite ethnicity and urban/rural covariate is the only explanatory variable of the model). The values of the coefficients for Asian/urban, Black/urban, Mixed/urban, and Nonwhite/rural are all higher in the model without covariates. This is due to the impact of other covariates not included in the simple regression model. For instance, individuals of Asian, Black, or Mixed ethnicity are typically younger than individuals of White ethnicity, and children and teenagers typically have more contacts, while retired individuals have fewer contacts on average.

**Figure S12: Value of ethnicity-related coefficients in two regression models: the full model presented in the main analysis, and a model with ethnicity and urban/rural status as only covariate.** Top panel: coefficients influencing the mean number of contacts. Bottom panel: coefficients influencing the dispersion of the number of contacts.

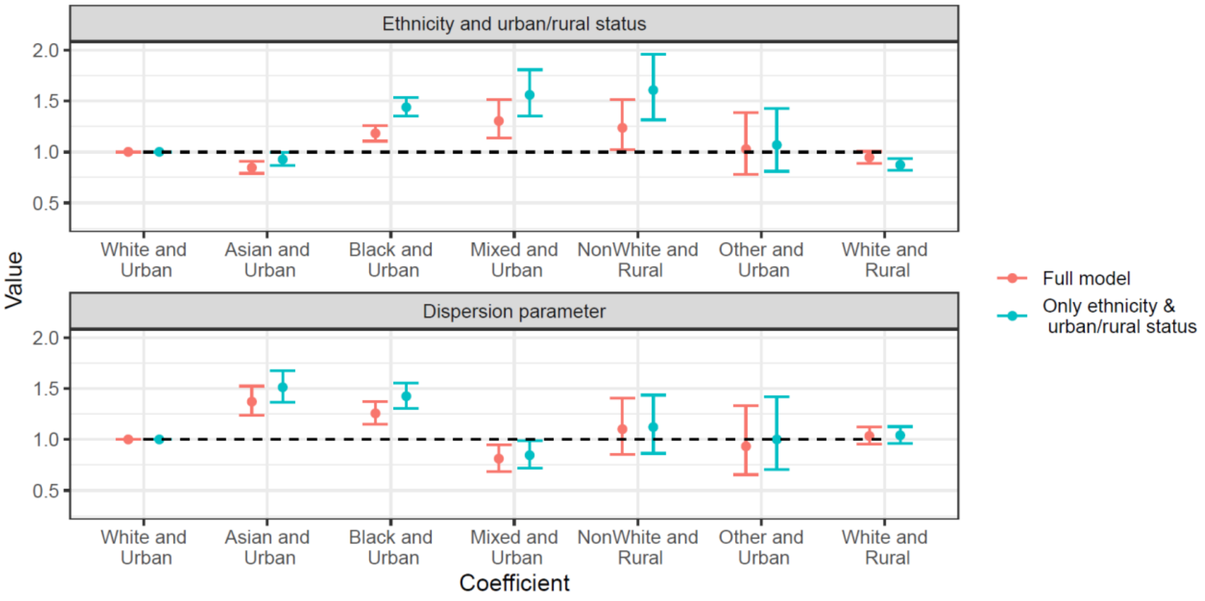


### Supplementary Section S7. Simulation of the distribution of contacts from regression outputs

In this section, we simulate the distribution of contacts per ethnicity in synthetic populations under a range of hypotheses to visualise the impact of the regression outputs and demographic characteristics. This is separate from the synthetic populations used to create contact groups, and from the transmission model. These simulations only show how the regression coefficients and the demographic characteristics of each ethnic group impact the distribution of contacts.

We created synthetic populations using the distribution of age by ethnicity, household size by age and ethnicity, annual income by ethnicity, and employment status by ethnicity, age, and gender in England. Since the 2021 Census only reports a binary “Male” / “Female” variable, “Other gender” was not included in the simulation. In each hypothesis, we generated 50 synthetic populations, and each synthetic population contained 10,000 individuals per ethnic group. The number of contacts were simulated using the Urban level for each ethnic group (i.e. Asian and Urban, Black and Urban, Mixed and Urban, Other and Urban, White and Urban)

First, we explored how ethnicity-related regression coefficients impact the distribution of contacts. To do so, we set all demographic variables of individuals in the synthetic population to their reference level in the regression model (Supplementary Figure S13, column A, top panel). In line with the regression coefficients, participants of Asian ethnicity had lower average number of contacts, and lower dispersion. In particular, the proportion of individuals of Asian ethnicity with more than 20 contacts was low (8% [95% Simulation Interval (SI): 6%-9%]), while the proportion of individuals of Mixed ethnicity with more than 20 contacts was 20% [95% SI: 18%-22%]. Also, the proportion of individuals of Mixed ethnicity with two or fewer contacts in the synthetic population was high (28% [95% SI: 26%-30%]), similar to the proportion in individuals of White ethnicity 29% [95% SI: 28%-30%].

Second, instead of setting all demographic variables to the reference levels from the regression model, we sampled the demographic characteristics of each individual in the synthetic population from the overall distribution in England according to the 2021 Census (Supplementary Figure S13, column A, middle panel). The overall mean number of contacts remained constant (10.3 [95% SI: 9.8-10.6] contacts), and the distribution by ethnicity was similar to the previous synthetic population, as the distribution of demographic characteristics was the same for all ethnicities.

Finally, we sampled the demographic characteristics of each individual from the ethnic-specific distribution in England according to the 2021 Census (Supplementary Figure S13, column A, bottom panel). The mean number of contacts in individuals of Mixed ethnicity increased (15.4 [95% SI 14.3-16.3], compared to 12.5 [95%SI 11.6-13.7] contacts in the previous hypothesis). The mean number of contacts in individuals of Asian ethnicity also increased (9.1 [95% SI 8.6-9.5], compared to 8.1 [95% SI 7.7-8.5] contacts in the previous hypothesis), becoming similar to the average in individuals of White ethnicity (9.2 [95% SI: 8.8-9.6]). The mean number of contacts in individuals of Black ethnicity remained high (12.1 [95% SI: 11.7-12.6]). The density plots of the 25% of individuals with the highest number of contacts per ethnicity showed that some simulated individuals of Mixed ethnicity reached 100 contacts, while the maximum number of simulated contacts for Asian and White individuals remained below 50 (Supplementary Figure S13, column B).

**Figure S13: Distribution of the number of contacts in the synthetic population by ethnicity, and density plot in the 25% of individuals with the highest number of contacts.** Top panels: synthetic population in which all individuals have the same level of demographic variables except for ethnicity. Middle panels: synthetic population in which the distribution of demographic variables for each individual is drawn from the national-level overall distribution. Bottom panels: synthetic population in which the distribution of demographic variables for each individual is drawn from the national-level distribution by ethnicity. The error bars correspond to the 95% simulation interval.

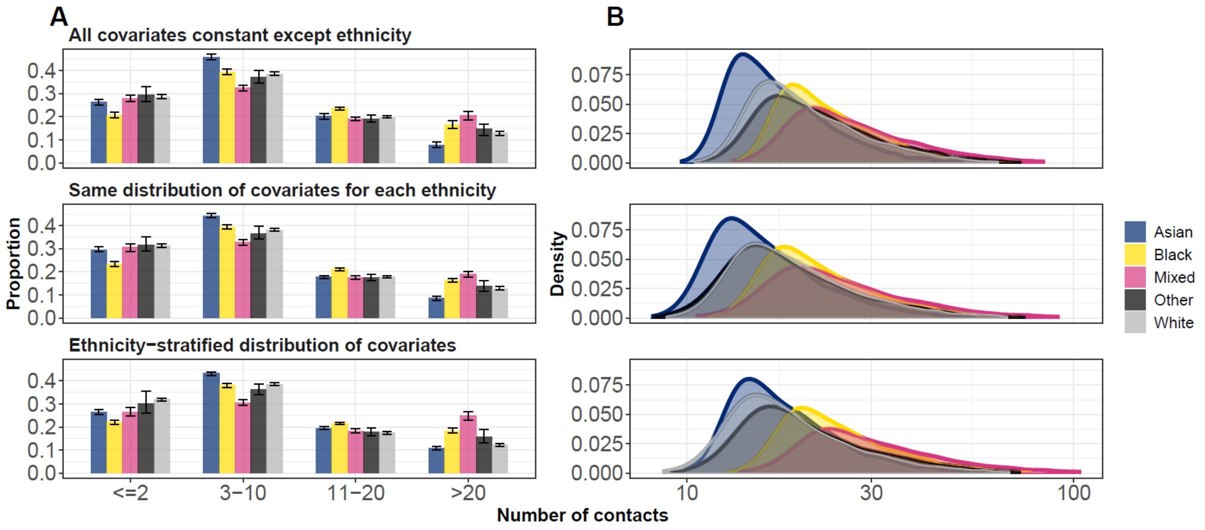


### Supplementary Section S8. Age-standardised attack rate

We computed the age-standardised attack rate and relative attack rates by ethnic group for each scenario (Supplementary Figure S14). Differences remain between ethnicities, but the age-standardised relative attack rates are closer to 1 than the raw relative attack rates in the “Representative of England census” scenario, and the scenario where we set ethnicity-related regression coefficients to 1 (Supplementary Figure S14, top panels, first two columns). In particular, the age-standardised attack rate is highest in individuals of Black ethnicity, as individuals of Mixed ethnicity are on average much younger than all other ethnic groups. In the other two scenarios, the age-standardised attack rates are the same as the raw attack rates since we used the same age structure in each ethnic group.

**Figure S14: Age-standardised transmission dynamics and sources of heterogeneity.** Top panels: age-standardised attack rate by ethnic group for each scenario and value of $R_0$. Bottom panels: Age-standardised relative attack rate compared to the White ethnic group for each scenario and value of $R_0$. The lines represent the median values, and the ribbons show the 95% Simulation intervals.

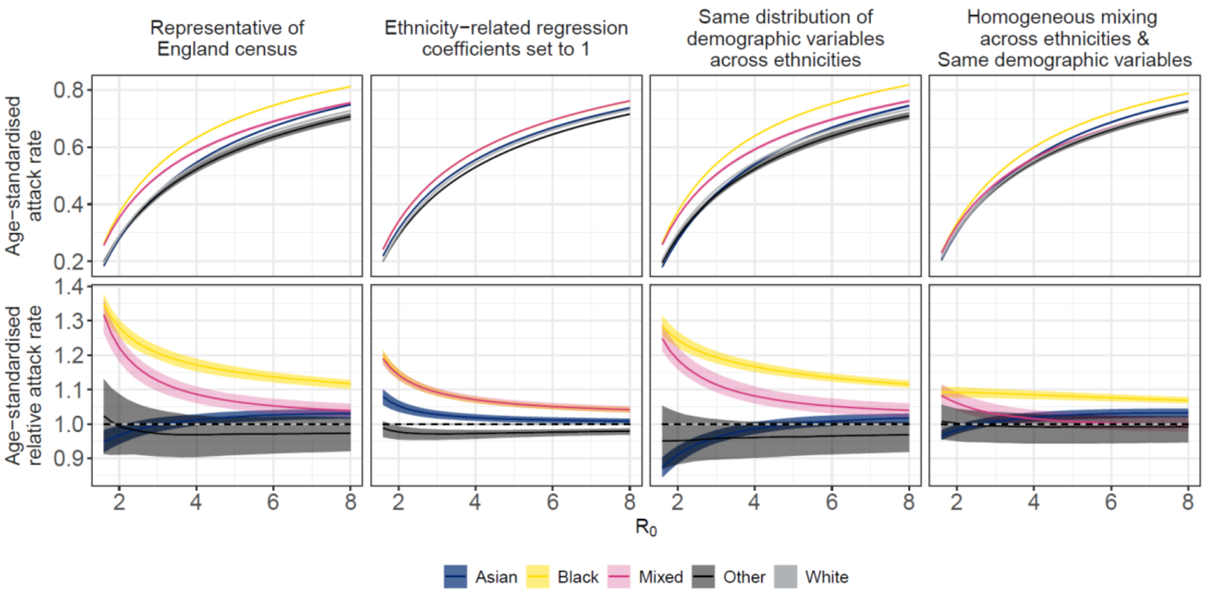


### Supplementary Section S9. Transmission dynamics

The model was implemented with three parameters: the duration of the infectious period (set to 5 days), the duration of the incubation period (set at 3 days), and the transmissibility rate. Changes to the incubation and infectious period would not impact the final size and relative attack rate between ethnicities. The model was run by either specifying $R_0$ (the basic reproduction number), or $\beta$ (the transmission rate). The population size depended on the simulation settings, and we set the initial number of exposed individuals to 30 in order to avoid epidemics dying out early due to stochasticity. All other individuals in the population started off as susceptible.

**Figure S15: Model trajectories in 10 simulations generated with $R_0$ = 2, stratified by age (top panels), ethnicity (middle panels), and contact group (bottom panels).** The left column shows the overall number of cases infected through the course of the simulated epidemics in each group. The middle column shows the attack rate per group through the course of the simulated epidemics.The right column shows the daily proportion of new cases in each group.

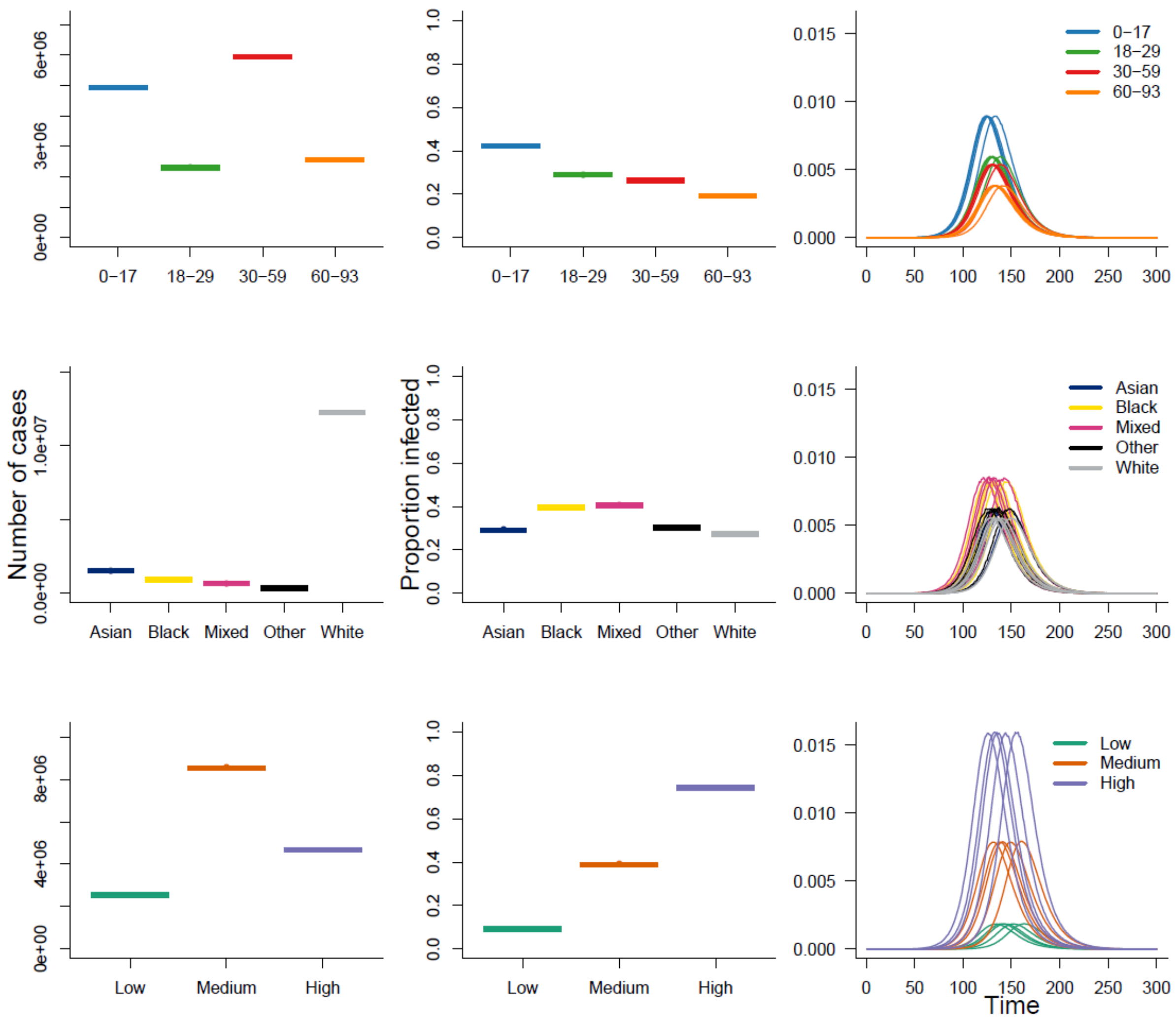

**Figure S16: Model trajectories in 10 simulations generated with $R_0$ = 4, stratified by age (top panels), ethnicity (middle panels), and contact group (bottom panels).** The left column shows the overall number of cases infected through the course of the simulated epidemics in each group. The middle column shows the attack rate by group through the course of the simulated epidemics.The right column shows the daily proportion of new cases in each group.

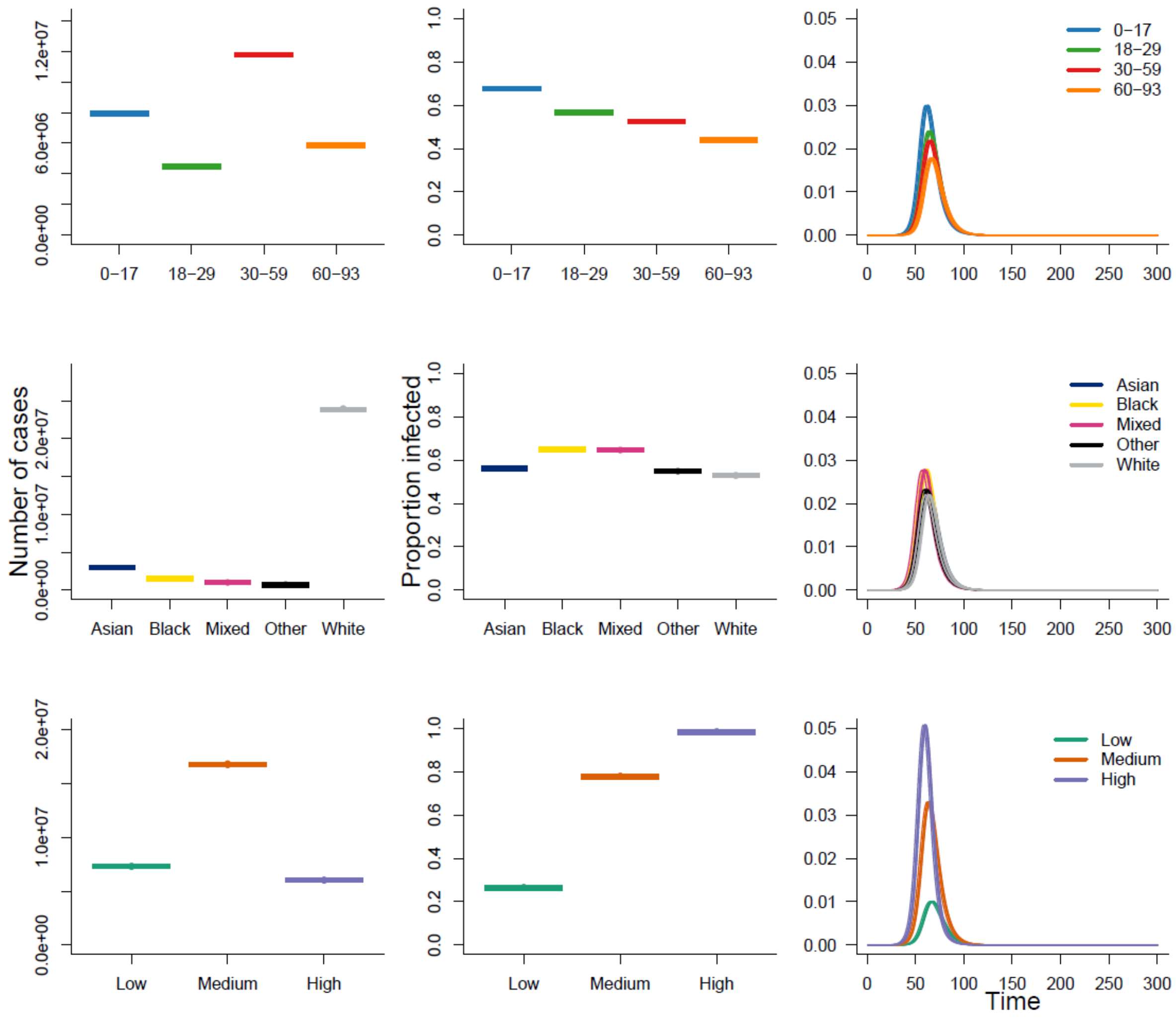


### Supplementary Section S10. Sensitivity analysis with four contact groups

As a sensitivity analysis, we ran the simulation analysis presented in Figure 3 with four contact groups instead of three. Adding a fourth contact group increased the overall amount of heterogeneity in the model, which led to a slight decrease in overall attack rate for a given value of $R_0$ (bottom panel of Supplementary Figure S17). When using data representative of the English census, the overall attack rate generated with $\beta = 0.00335$ is now higher than that obtained with $R_0$ = 4. This is in line with the fact that positively correlated additional heterogeneity at fixed parameters increases $R_0$ (3). However, adding a fourth group did not change the relative attack rate between ethnic groups.

**Figure S17: Simulated transmission dynamics and sources of heterogeneity with four transmission groups.** Top panels: Proportion of each ethnic group infected across the course of the epidemics for each scenario and value of $R_0$. Middle panels: Relative attack rate compared to the White ethnic group for each scenario and value of $R_0$. The lines represent the median values, and the ribbons show the 95% Simulation intervals. Bottom panel: Density of the overall proportion of the population infected for each scenario, with $R_0 = 4$, or $\beta = 0.0335$ (which corresponds to $R_0 \sim 4$ in the reference scenario with three contact groups).

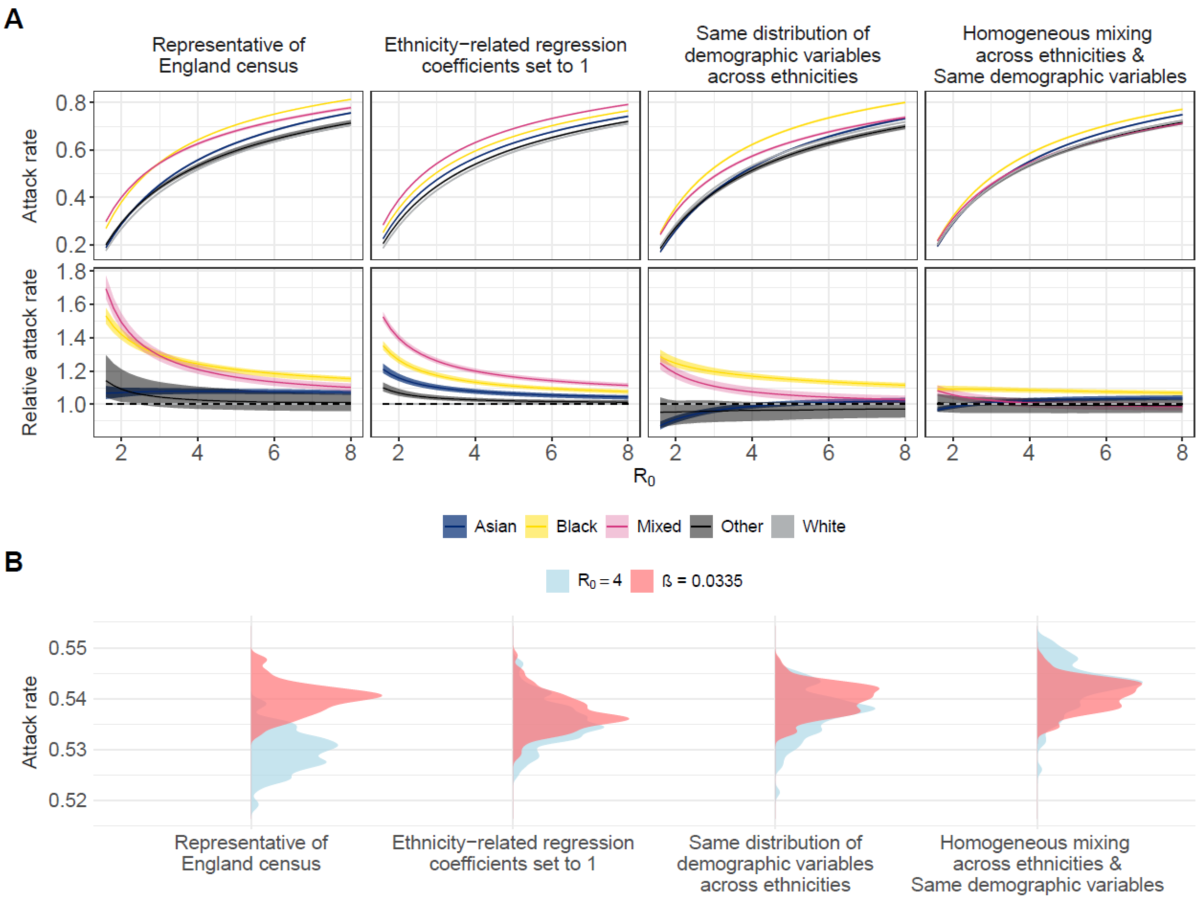


### Supplementary Section S11. Demography by city

We quantified the impact of local demographics by generating stochastic simulations using the population characteristics of six urban centres in England: Birmingham, Leicester, Liverpool, London, Manchester, and York. These cities were selected as they capture different distributions of ethnicity: Liverpool and York have a comparatively high proportion of individuals of White ethnicity (84% and 94%, respectively), the share of individuals of Asian ethnicity in Birmingham and Leicester are among the highest in England (31% and 44% respectively), while Manchester and the region of London have high proportions of individuals of Black ethnicity (12% and 14%, respectively). We adapted the per capita age and ethnicity transmission matrices from Reconnect to each city using the density correction approach described by Arregui et al (2), whereby the contact rate in the new population is equal to the contact rate in the reference population multiplied by the ratio of the reference to the new population size.

**Figure S18: Demographic characteristics of the population in Birmingham and Leicester.** Cumulative age distribution of the population in Birmingham and Leicester, stratified by ethnicity, with the pie chart insert showing the distribution by ethnicity in the two cities (from 2021 Census data). Bar charts: distribution of household size and employment status by ethnicity in the two cities (from 2021 Census data).

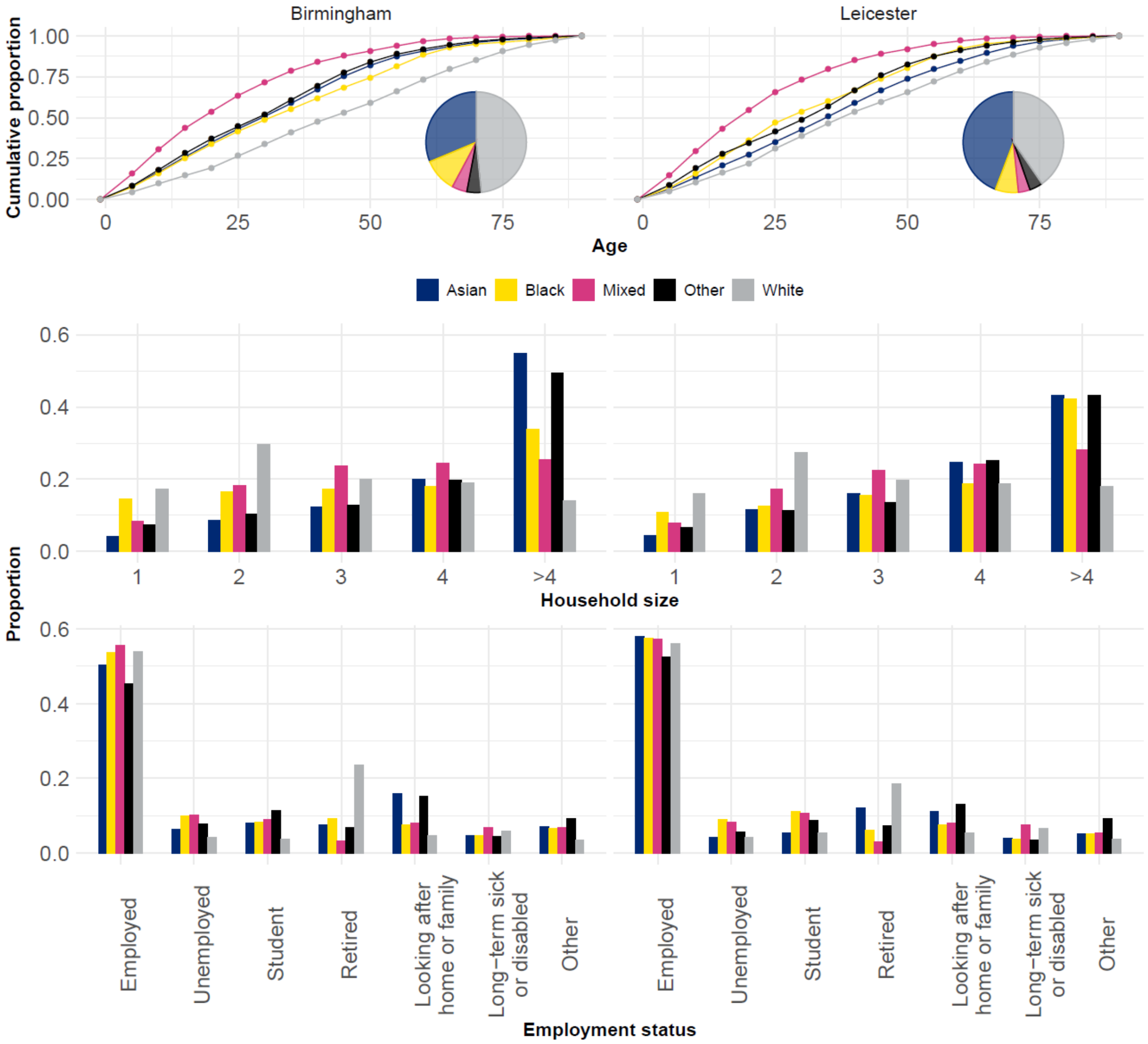

**Figure S19: Demographic characteristics of the population in London and Manchester.** Cumulative age distribution of the population in London and Manchester, stratified by ethnicity, with the pie chart insert showing the distribution by ethnicity in the two cities (from 2021 Census data). Bar charts: distribution of household size and employment status by ethnicity in the two cities (from 2021 Census data).

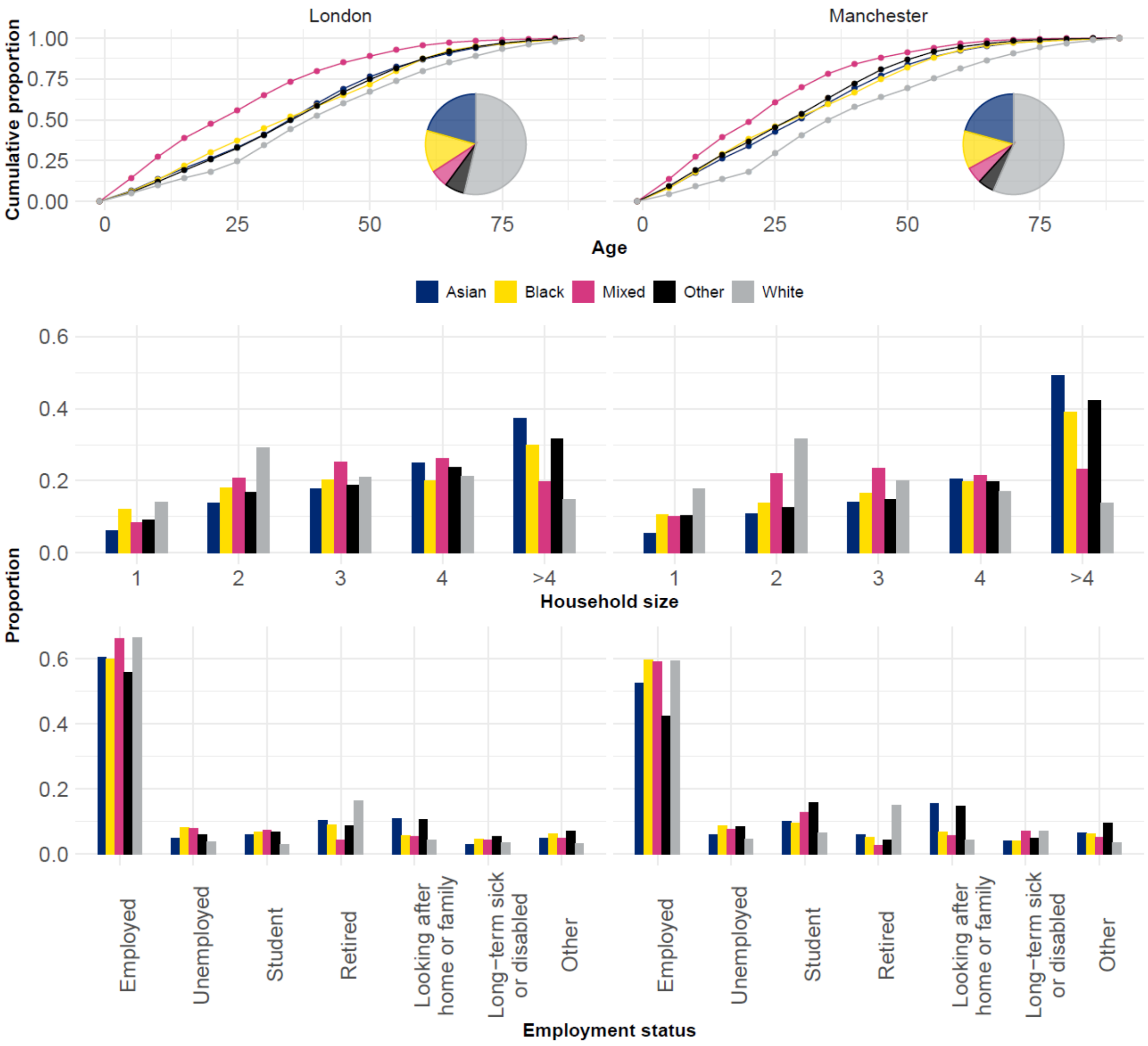

**Figure S20: Demographic characteristics of the population in Liverpool and York.** Cumulative age distribution of the population in Liverpool and York, with the pie chart insert showing the distribution by ethnicity in the two cities (from 2021 Census data). Bar charts: distribution of household size and employment status by ethnicity in the two cities (from 2021 Census data).

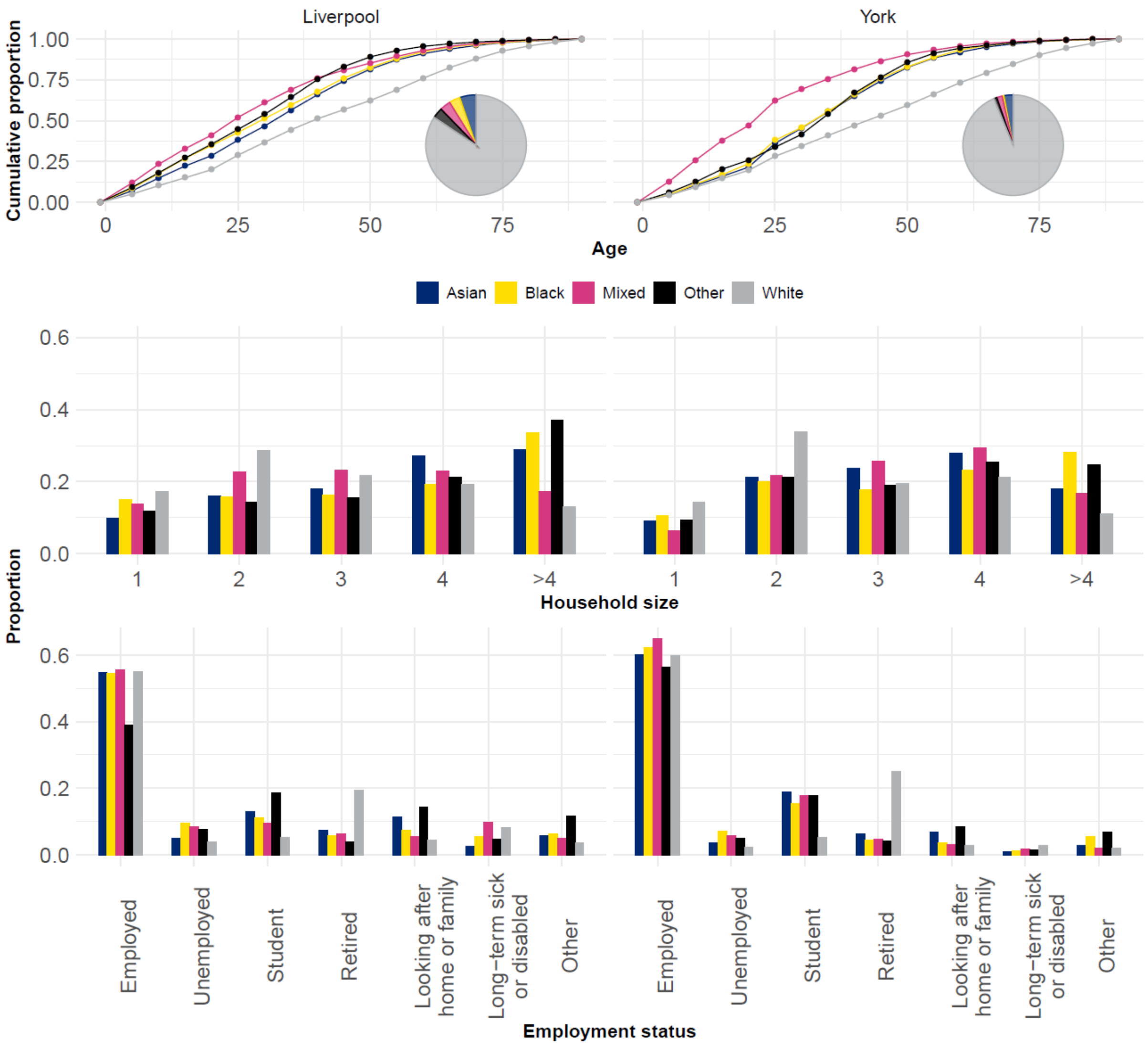


### Supplementary Section S12. Attack rate by city

The attack rate by ethnicity highlights how local distribution of age, ethnicity, household size, and employment status impact simulated outbreak dynamics (Figure S21). The overall attack rate was lower in York and Liverpool across all values of $R_0$, but the attack rate in the White ethnic group was lower in Birmingham, Leicester, London, and Manchester.

**Figure S21: Attack rate by ethnic group for each city and value of** R$_0$**.** The lines represent the median values, and the ribbons show the 95% Simulation intervals.

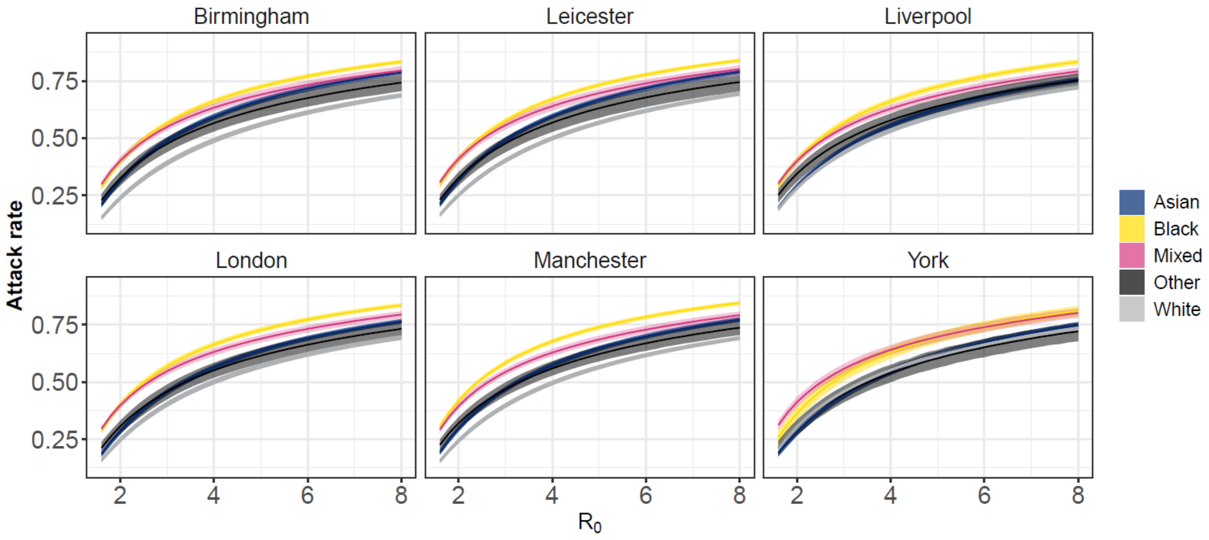


Across all cities, individuals of Black ethnicity had the highest age-standardised attack rate (Figure S22). The age-standardised attack rate in individuals of Mixed ethnicities was lower than the raw attack rates, which is expected, since individuals of Mixed ethnicities have a younger age distribution relative to other ethnicities.

**Figure S22: Age-standardised relative attack rate of each ethnic group compared to the White ethnic group across the course of the epidemics for each city and value of** $R_0$**.** The lines represent the median values, and the ribbons show the 95% Simulation intervals.

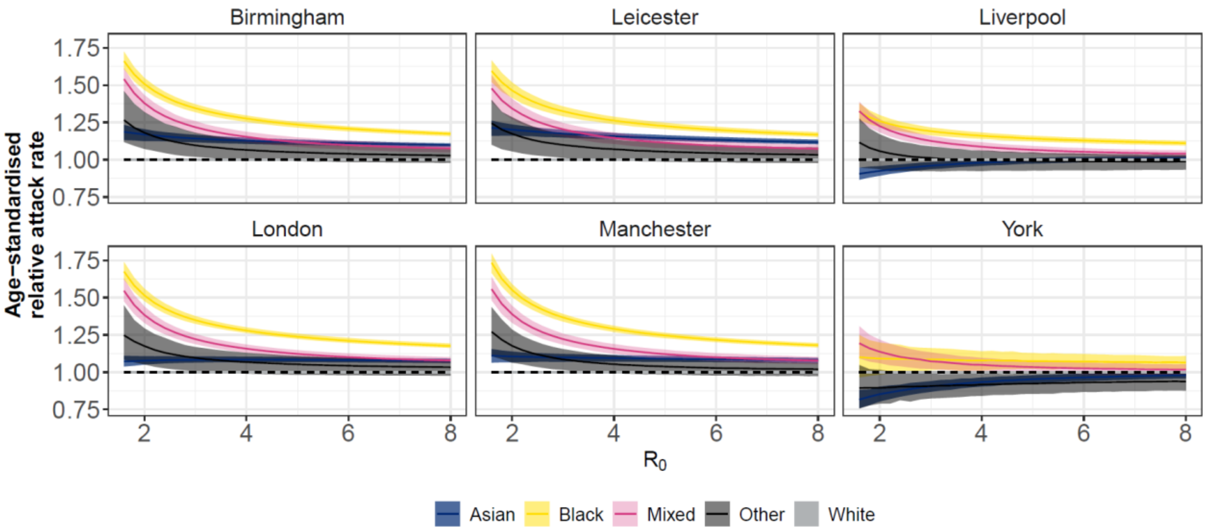
